# Trends and inequalities in statin use for the primary and secondary prevention of cardiovascular disease between 2009 and 2021 in England

**DOI:** 10.1101/2024.11.22.24317782

**Authors:** Rutendo Muzambi, Krishnan Bhaskaran, Helen Strongman, Tjeerd van Staa, Liam Smeeth, Emily Herrett

**Affiliations:** Faculty of Epidemiology and Population Health, London School of Hygiene and Tropical Medicine, London, WC1E 7HT, UK; Faculty of Epidemiology and Biostatistics, School of Public Health, Imperial College London, London, W12 0BZ, UK; Centre for Health Informatics, Division of Informatics, Imaging and Data Science, Faculty of Biology, Medicine and Health, School of Health Sciences, The University of Manchester, Manchester Academic Health Science Centre, Vaughan House, Manchester, M13 9PL, UK

## Abstract

**Objective:** To investigate trends and inequalities in statin use for the primary and secondary prevention of cardiovascular disease (CVD)

**Design:** Repeated cross-sectional and historical cohort study designs

**Setting:** English primary care electronic health records from the Clinical Practice Research Datalink (CPRD Aurum) linked to Hospital Episode Statistics Admitted Patient Care

**Participants:** 5 million adults aged 25 years and older randomly sampled from CPRD Aurum between 1^st^ April 2009 and 31^st^ December 2021.

**Outcome measures:** Monthly proportion of current statin users; adjusted odds ratios (aOR) for statin initiation; adjusted hazard ratios (aHR) for cardiovascular risk assessment, statin discontinuation and statin re-initiation and number of CVD events prevented with optimal statin use and estimated costs saved.

**Results:** The overall monthly proportion of individuals prescribed statins for primary prevention increased from 22.3% in 2009 to 35.6% in 2021 among those aged 70+ years, and was stable in other age groups. The proportion of eligible individuals receiving a statin for secondary prevention was higher in all age groups (e.g. increasing from 68.1% to 73.7% over the same period, in those aged 70+). Overall prevalence of statin use was lowest among women, 25-39 age group, and black, mixed, and other ethnic groups for both primary and secondary prevention. Monthly proportion of CVD risk assessment, among those eligible, increased from 13.7% in May 2009 to 31.8% by November 2021. 79.0% of individuals were initiated statins within 60 days of a CVD event. Women (aOR 0.70; 95% CI, 0.68 − 0.72) and people of black ethnicity (aOR 0.71; 95% CI, 0.65-0.77) were less likely to be initiated statins compared to those of white ethnicity while people of south Asian ethnicity (aOR 1.53; 95% CI, 1.42-1.64) were more likely to be initiated statins than white people for secondary prevention. Statin discontinuation was most likely among women (aHR 1.08, 95% CI; 1.06−1.11) black people (aHR 1.76, 95% CI, 1.65−1.89) and the most deprived group (aHR 1.08, 95% CI; 1.04−1.12) compared to men, white people and the least deprived group, respectively, for primary prevention with similar associations seen for secondary prevention for ethnicity and deprivation. With optimal statin treatment, over 150,000 cardiovascular events could be prevented in the next 10 years for primary prevention and 5 years for secondary prevention resulting in a potential saving to the health service of over £400 million in those eligible for statins.

**Conclusion:** Statin use remains suboptimal and inequalities particularly among women, people of black ethnicity and those in the most deprived socioeconomic groups persist across multiple stages of statin use for both primary and secondary prevention. To reduce these inequalities and avoid missed opportunities to prevent cardiovascular events and costs to the NHS, strategies are needed specifically targeting these patient groups to reduce the burden of CVD.

**Summary box:** *What is already known on this topic:*

- Previous studies have shown that statins are under-prescribed and under-used in both primary and secondary prevention of cardiovascular disease (CVD) resulting in missed opportunities to reduce CVD burden.
- Few studies have examined trends in statin use in recent years, including during the pandemic period.
- Inequalities in statin use based on age, gender, ethnicity and deprivation have been identified previously, however it is unclear where, along the pathway from identification of eligible patients to initiation and continuation of statins, these inequalities manifest.

*What this study adds:*

- Levels of CVD risk assessment were suboptimal throughout the study period: a modest increase in the proportion of eligible individuals with a CVD risk assessment from 14% in May 2009 to 35% in February 2020 was followed by a decline during the COVID-19 pandemic.
- Prevalence of statin use increased between 2009 and 2021 but remained suboptimal throughout, and with important sociodemographic disparities. Women and people of black ethnicity were less likely to initiate statins for secondary prevention compared to men and the white ethnic group while people of south Asian ethnicity and the 60-69 age group were more likely to initiate statins compared to people of white ethnicity and the 25-39 age group, respectively.
- Statin discontinuation was higher among women than men (secondary prevention only), black ethnic groups compared to white ethnic groups, and the most deprived socioeconomic groups compared to the least deprived for both primary and secondary prevention.
- We estimated that over 100,000 cardiovascular events could be prevented in the next 10 years if the observed missed opportunities for statin use in primary CVD prevention among eligible individuals were fully addressed, and a further 50,000 events could be prevented over 5 years for secondary prevention. The consequent potential saving to the health service was estimated to be over £400 million.

**Summary of main results:** 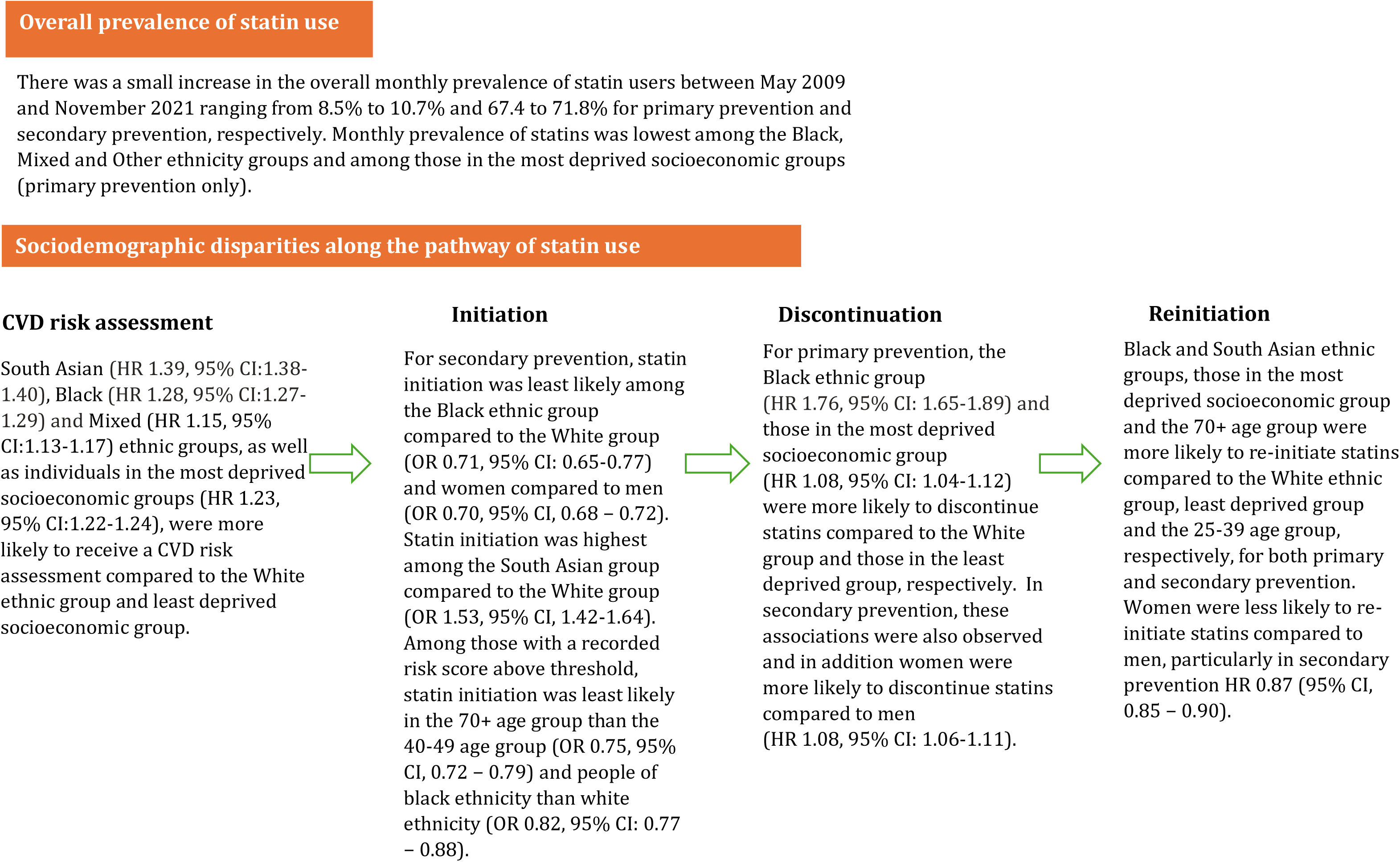

## Introduction

Cardiovascular disease (CVD) is the leading cause of morbidity and mortality worldwide, accounting for 32% of all global deaths in 2019 and a quarter of all deaths in the UK.(1, 2) Statins have been proven to be effective in reducing cardiovascular events and are widely prescribed, with atorvastatin the most commonly dispensed medication in England between 2022 and 2023. (3–5)

Despite National Institute for Health and Care Excellence (NICE) guidelines recommending the use of statins in those at risk of CVD and to all individuals with a prior CVD event, evidence suggests that statins are under-prescribed, with previous studies showing that only 30-50% of adults at high risk of CVD in the UK were receiving statins.(6–9) Coverage of CVD risk assessment is low and studies investigating trends in CVD risk assessment in more recent years including during the COVID-19 pandemic are lacking and evidence on inequalities in risk assessment is limited.(10)

Disparities in statin prescribing in terms of age, gender, ethnicity and deprivation have been highlighted previously.(7, 11–17) However, it is unclear where along the pathway from identification of eligible patients, statin initiation and continued statin use these inequalities manifest. Missed opportunities for treatment are associated with worsening of clinical outcomes, mortality and higher healthcare costs.(14, 18, 19)

Therefore, we examined trends and factors (age, gender, ethnicity, and deprivation) associated with statin prevalence, cardiovascular risk scoring, initiation (among those with an above threshold CVD risk score for primary prevention or existing CVD event for secondary prevention), discontinuation, and re-initiation of statins between 1^st^ April 2009 and 31^st^ December 2021. We also estimated the number of cardiovascular events that could be prevented and healthcare cost savings with optimal statin use.

## Methods

### Data and participants

We conducted a several cross-sectional and historical cohort studies using data from the Clinical Practice Research Datalink (CPRD Aurum) linked to Hospital Episode Statistics (HES) Admitted Patient Care.(20, 21) CPRD Aurum database consists of pseudonymous electronic health records from UK primary care practices using the EMIS general practice (GP) electronic health record software. The CPRD Aurum January 2022 build was used to extract the cohort for this study. CPRD Aurum contained over 40 million individuals and 1,489 GP practices.(22) The database includes information on patient and practice demographics, diagnoses and symptoms, medication prescriptions, and referrals. Our study followed the Reporting of Studies Conducted Using Observational Routinely Collected Data reporting guidelines.(23)

Our overall study population included 16,650,277 individuals aged 25 years and older between 1st April 2009 and 31st December 2021 eligible for HES linkage and at least 12 months research standard follow up in CPRD Aurum (Supplementary Figure 1). From this population, we extracted data from a random sample of 5 million individuals and details of the random sample can be found in the Supplementary Methods and Supplementary Table 1. Our study period was from April 2009 as this corresponds to when NHS health checks, which assess CVD risk and can inform statin prescribing, were first introduced.(24) Start of follow up for the overall cohort was defined as the latest of 1^st^ April 2009, 25^th^ birthday and 12 months after current registration in the CPRD practice. End of follow-up was the earliest of 31^st^ December 2021, death date or end of follow-up in CPRD.

We used SNOMED codes in CPRD and ICD-10 codes in HES to identify CVD diagnoses and covariates. Statin prescription information was obtained in CPRD using Dictionary of Medicines and Devices (dm+d) codes.

Figure 1 shows a conceptual figure of the pathway of statin use. The overall prevalence of statin use for primary and secondary prevention is the result of identification of eligible individuals, initiation and continued use of statins. Our study seeks to determine where, and in whom, along this pathway are there inequalities in statin treatment and thus are opportunities for CVD prevention being missed.

**Figure 1.**
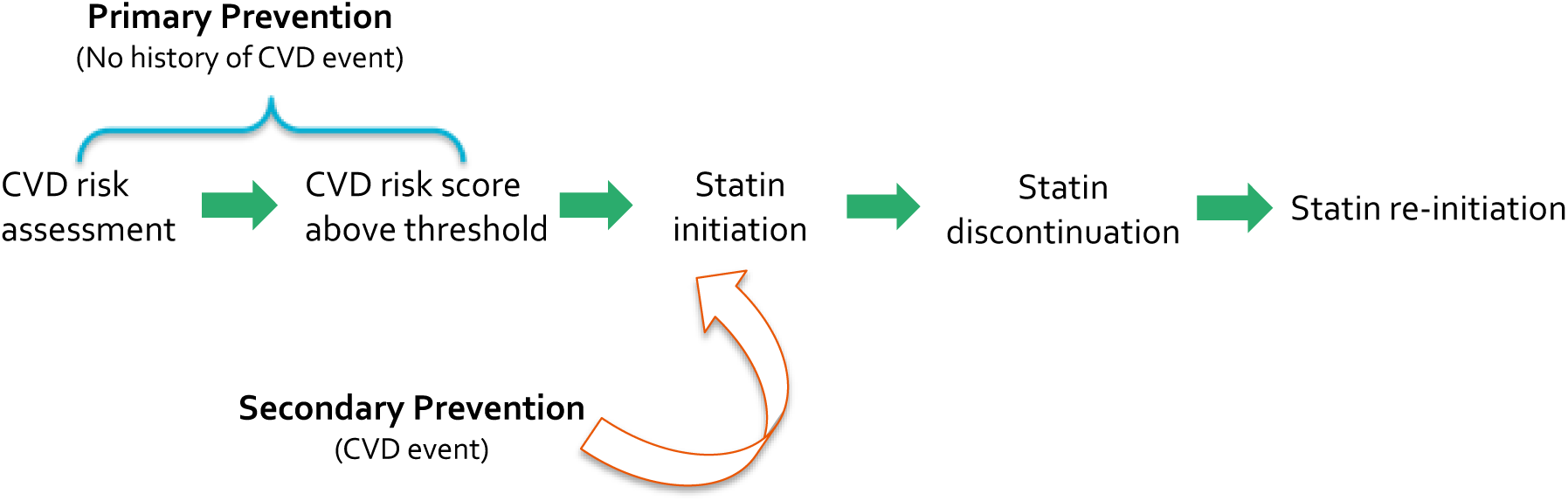
Conceptual figure depicting typical pathway of statin use

Our subpopulation cohorts are described below.

### Study population and statistical analyses

#### CVD risk scoring

We included individuals without a CVD diagnosis aged between 25-84 years old. We described the characteristics of individuals formally risk scored by their GP using all available CVD risk assessment tools (QRISK, QRISK2, Framingham, Joint British Society, ASSIGN, or unspecified).(25–28) Then, we described the monthly proportion of individuals with a recorded cardiovascular risk score in the last 5 years, between May 2009 and December 2021, and stratified by CVD risk tool, CVD risk category as assessed by the GP (<10%, 10-19% and 20%), age, gender, ethnicity, and deprivation. Individuals were excluded from a monthly denominator when they were diagnosed with a CVD event, became ineligible for CVD risk assessment (e.g. CKD, T1DM or familial hypercholesterolaemia diagnosis) or were initiated on statins. Individuals were defined as having a current CVD risk record if they had a risk record in the last 5 years (corresponding to the recommended frequency of CVD risk assessment).(24) If individuals had multiple CVD risk scores on the same date, we kept the highest score.

To determine which individuals were eligible for CVD risk assessment, we included individuals aged 40-74 years (in accordance with the NHS health check age criteria), with no history of CVD, chronic kidney disease (CKD), type 1 diabetes mellitus (T1DM), familial hypercholesterolaemia and no contraindications to statins. Cox proportional hazards regression models, with the start of follow up as the time scale, were used to estimate hazard ratios (HRs) for the association between demographic variables (age at start of follow up, gender, ethnicity, and deprivation) and receiving a CVD risk score from the GP. Start of follow up was defined as the latest of 1^st^ April 2009, 40^th^ birthday and 12 months after current registration in CPRD. Individuals were followed until the earliest of CVD risk assessment, 75^th^ birthday, CVD event or ineligibility for CVD risk assessment. We adjusted for demographic variables mentioned above as these factors are likely to be fixed. We had insufficient time-updated information to adjust for time-dependent variables.

### Statin prevalence

To describe the prevalence of statin use for primary CVD prevention, we included individuals without prior CVD (e.g. past or current history of myocardial infarction, angina, revascularisation procedures, stroke, transient ischaemic attack, or peripheral arterial disease). For secondary prevention, our cohort included individuals with existing CVD. We excluded individuals contraindicated to statins (using SNOMED codes for liver disease or codes explicitly stating statin contraindication) in both cohorts. An individual was defined as a current statin user if they had an ongoing statin prescription based on the number of days prescribed, allowing for a grace period of 28 days between prescriptions.

We described the proportion of individuals with a current statin prescription each calendar month for primary and secondary prevention. In subgroup analyses, monthly prevalence was stratified by age (25-39, 40-49, 50-59, 60-69, 70-79, 80+), gender (men and women), ethnicity, (white, South Asian, black, mixed and other), deprivation (using linked patient-level Townsend deprivation scores, categorised into quintiles) and CVD subtype (for secondary prevention). Individuals left both prevalence cohorts when they had a statin contraindication and left the primary prevention cohort when they were diagnosed with a CVD event.

### Statin initiation

To assess initiation of statins for primary prevention, we included individuals aged 40-74 who were eligible for the NHS health check and had a recorded CVD risk score above the threshold for statin initiation, with no history of CVD, CKD, T1DM or familial hypercholesterolaemia. We included the first recorded CVD risk score, within follow-up, that was eligible for statin initiation where there was no previous statin prescription.

Consistent with NICE guidelines, those with a 20% or higher 10-year risk of CVD before 2014 and a 10% or higher 10-year risk of CVD after 2014, were considered above the treatment threshold and eligible for statin initiation.(29) Statin initiation was defined as having a first statin prescription within 28 days of a first recorded CVD risk score above treatment threshold.

Our secondary prevention initiation cohort comprised individuals with a CVD event who remained alive and under follow-up within 60 days of a CVD event and with no statin contraindications. Statin initiation was defined as having a first statin prescription within 60 days of a CVD event. We chose a 60-day grace period between the first cardiovascular event and subsequent statin prescription to account for individuals who may be initiated statins in secondary care and may wait until they have completed their prescription before obtaining a statin prescription in primary care.

We used Kaplan Meier curves to examine the time from first CVD risk score above threshold to statin initiation (primary prevention) or time from CVD event to statin initiation (secondary prevention). These curves were stratified by age, gender, ethnicity, deprivation, CVD risk score (primary prevention only) or CVD subtype (secondary prevention only). Individuals were censored at end of follow up (including death), CVD event (primary prevention only) and statin contraindication (e.g. liver disease). We used logistic regression to examine the factors associated with initiating a statin for primary and secondary prevention within 28 days of becoming eligible. We adjusted for age at risk assessment (primary prevention) or age at CVD diagnosis (secondary prevention), gender, ethnicity, deprivation, Body Mass Index (BMI, kg/m^2^) (underweight, normal weight, overweight, obese or morbidly obese), smoking status (non-smoker, current smoker and former smoker), treated hypertension (defined as antihypertensives prescribed on the date of or after hypertension diagnosis), type II diabetes mellitus (T2DM), atrial fibrillation and rheumatoid arthritis. We did not adjust for HDL, LDL, total cholesterol, and systolic and diastolic blood pressure due to moderate/high levels of missingness (Supplementary Table 2) and chronic kidney disease (in primary prevention analyses) as CVD risk assessment is not recommended in this population for primary prevention.

### Statin discontinuation

We included all individuals aged 25 years and older who had initiated statins for primary prevention (all individuals without pre-existing CVD event) and secondary prevention (all individuals with a first CVD event during the study period). After individuals reached the end of their prescription based on the date of prescription, dose and quantity prescribed, statin cessation was defined as reaching the end of a 90-day grace period without a new statin prescription, to allow time for those with overlapping prescriptions to use their excess supply.

We used Kaplan Meier curves to examine the time from statin initiation to discontinuation stratified by age, gender, ethnicity, deprivation, CVD risk category (primary prevention) or CVD subtype (secondary prevention). Cox proportional hazards regression models, with date of statin initiation as the time scale, were used to estimate hazard ratios (HRs) for the association between demographic variables (age at statin initiation, gender, ethnicity, and deprivation) and statin discontinuation, adjusted for these demographic variables.

### Statin re-initiation

Our re-initiation cohort included all individuals who had discontinued statins based on criteria mentioned above. For our primary prevention cohort, we additionally excluded individuals with a CVD diagnosis prior to statin cessation. Statin re-initiation was defined as receiving a subsequent statin prescription after a 90-day period without a statin prescription, starting from the time when the last prescription ran out (estimated based on date of previous prescription, dose and quantity of tablets prescribed).

We used Kaplan Meier curves to examine time from statin discontinuation to first subsequent statin prescription. We stratified by age at discontinuation date, gender, ethnicity, deprivation, CVD risk category or CVD subtype. We used Cox proportional hazards regression models, with date of statin discontinuation as the time scale, to estimate hazard ratios (HRs) for the association of age, gender, ethnicity, and deprivation with statin discontinuation, adjusting for these demographic variables.

### Healthcare costs

We used the National Schedule of NHS (National Health Service) costs to calculate the cost of CVD events for the year of 2021-22 for all NHS trusts and NHS foundation trusts and all healthcare resource groups (Supplementary Table 3). (30) We also used the prescription cost analysis dataset for the financial year 2021-22 which includes the costs of medications dispensed in the community in England (Supplementary Table 4).(31)

To calculate the number of additional cardiovascular events avoided over 10 years with optimal statin use for each risk category, we calculated the 10-year total number of expected events from the CVD risk, based on the recorded risk scores among those with a CVD risk assessment, and number of individuals not taking statins, and applied a 25% risk reduction due to statins for those without vascular disease.(32) For secondary prevention, we calculated the number of cardiovascular events avoided over 5 years with optimal statin use for each CVD event among those eligible but not taking statins, using a 10% absolute benefit.(4) We then scaled these analyses to the English population using data from the 2021 census of adults aged 25 years and older (N=40,017,720).(33) We calculated the total cost of recommended statin dose for primary and secondary prevention (atorvastatin 20mg and atorvastatin 80mg) according to the British National Formulary using the total population not prescribed statins, total quantity, and cost per quantity from the 2021/22 English prescription cost analysis dataset. (31)

### Additional secondary and sensitivity analyses

We performed a range of secondary and sensitivity analyses. First, we stratified our initiation, discontinuation, and re-initiation analyses by the pre-pandemic (before 01/03/2020) and pandemic period (from 01/03/2020) to examine the effect of the COVID-19 pandemic. Second, we expanded our definition for statin initiation from a first statin prescription within 28 days after above threshold risk score to 90 days for primary prevention and from a statin prescription within 60 days after first CVD event to 90 days for secondary prevention. The rationale for this was to minimise misclassification of statin initiators. Third, to minimise misclassification of statin discontinuation, we expanded our definition of cessation from a 90-day grace period to 180 days.

### Model Checking

We found evidence of non-proportionality when we tested the Cox proportional hazards assumption using Schoenfeld residuals test and log-log plots for our analyses on factors associated with receiving a CVD risk score, statin discontinuation and statin re-initiation. To deal with this, we performed sensitivity analyses in which we added interaction terms between non-proportional demographic factors and time.

All analyses were conducted in R Studio, version 4.3.2. Numerical estimates of monthly proportions of statin users and individuals with CVD risk assessments as well as number of people at risk and events for the Kaplan Meier plots can be found on Github: https://github.com/RutendoMuzambi/trends_statin_use.

### Patient and Public Involvement Statement

No patients were involved in setting the research question or outcome measures, or the design or implementation of this study.

## Results

In our random subsample of 5 million individuals aged 25 years and older from a total population of 16.6 million individuals eligible for inclusion into the present study (Supplementary Figures 2 and 3), 4,606,274 and 416,431 individuals were included in the primary and secondary prevention cohorts specifically on statin use, respectively (Table 1A and 1B). Overall, individuals included in the primary prevention cohort were younger (median: 39.0 years [IQR, 29.0 to 53.0] vs 70.0 years [IQR, 59.0 to 79.0] and there was a higher proportion of women (50.8% vs 43.1%) than in the secondary prevention cohorts. We also present characteristics of individuals included in four sub-cohorts for prevalence, initiation, discontinuation, and re-initiation analyses in Table 1A and 1B. Of the 303,292 and 122,743 people in total who initiated statins during the study, 116,468 (38.4%) and 27,257 (22.2%) discontinued statins for primary and secondary prevention respectively (Supplementary Figures 2 and 3). Of these, 115,043 (79.5%) and 24,876 (91.3%) re-initiated statins for primary and secondary prevention respectively. Baseline characteristics of individuals included in the CVD risk assessment sub-cohorts on monthly prevalence of individuals with a CVD risk assessment and factors associated with a CVD risk assessment are presented in Supplementary Table 5.

**Table 1A:**
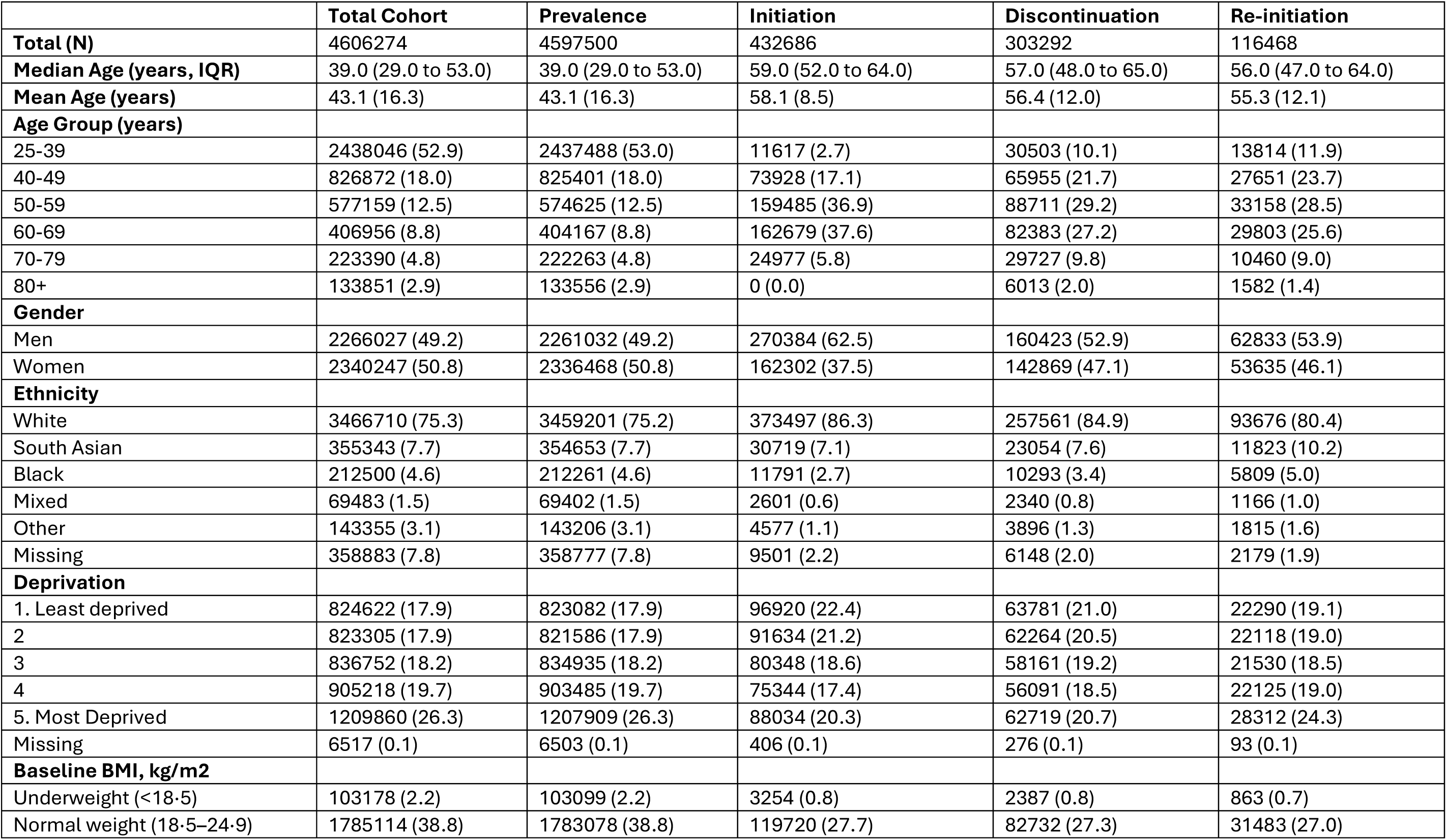
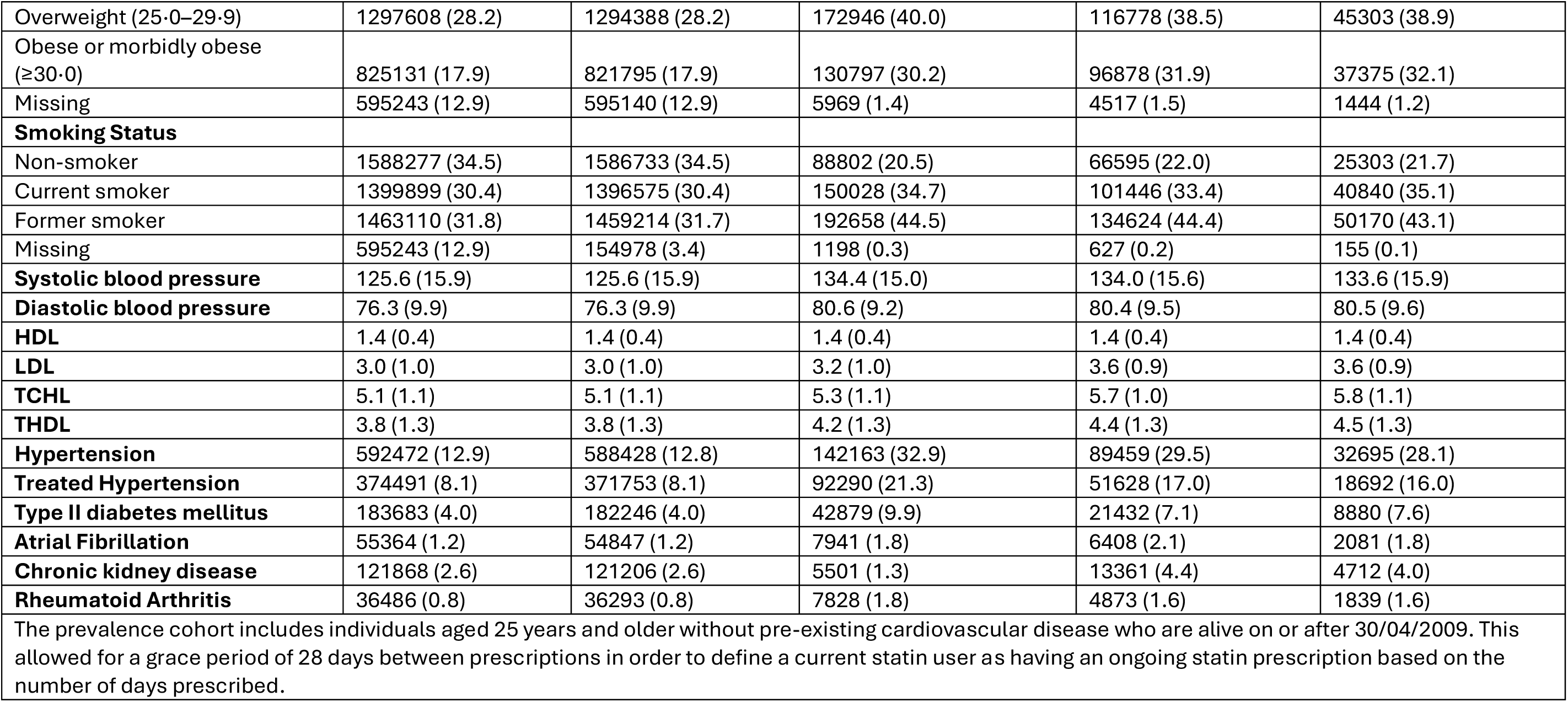
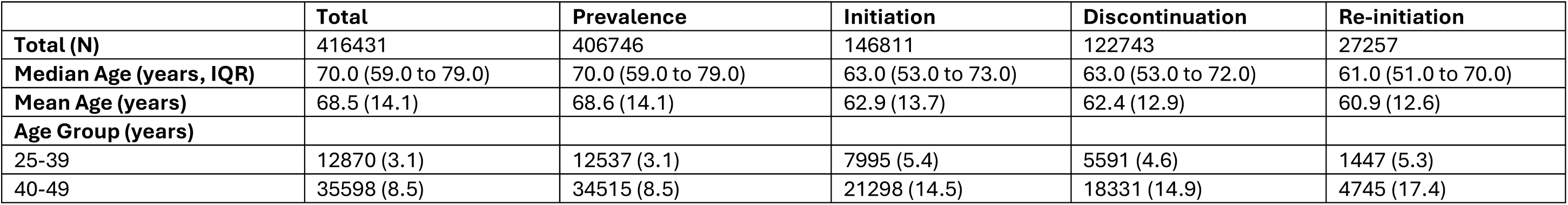
Baseline Characteristics of individuals included in the prevalence, initiation, discontinuation and re-initiation cohorts for primary prevention.

**Table 1B:**
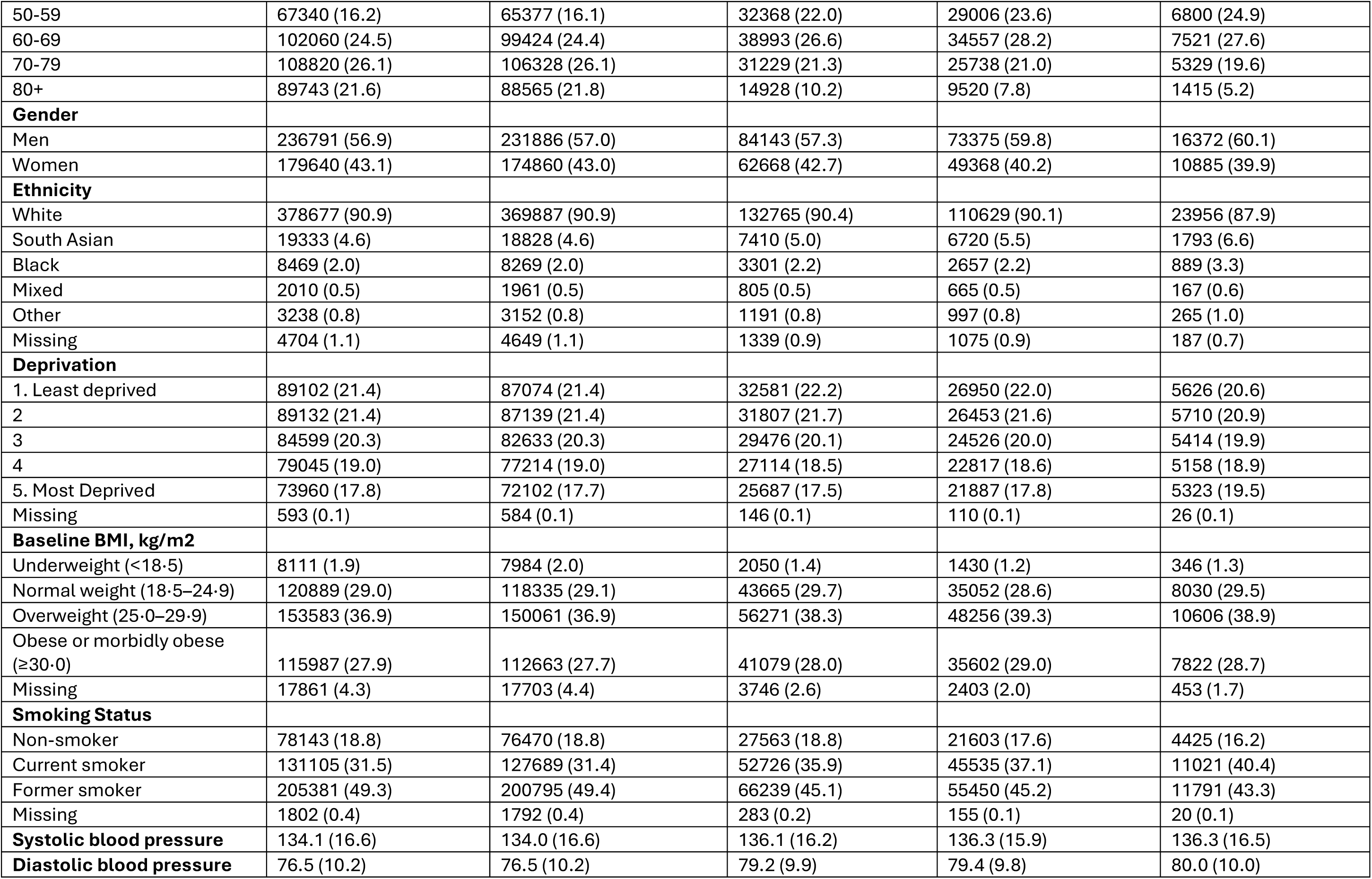
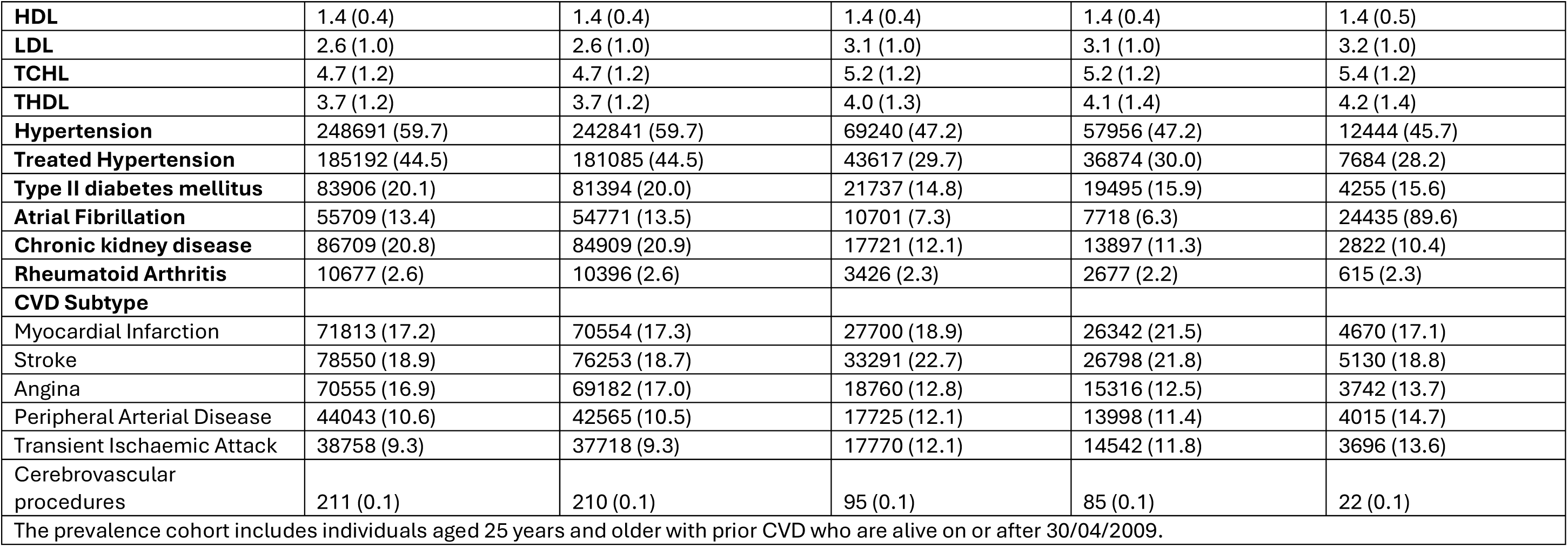
Baseline Characteristics of individuals included in the prevalence, initiation, discontinuation and re-initiation cohorts for secondary prevention.

Between May 2009 and November 2021, there was a small increase in the monthly proportion of statin users over time, ranging from 8.5% - 10.7% and 67.4-71.8% for primary prevention and secondary prevention, respectively (Figure 2). The increase in statin prevalence for primary prevention appears to be driven largely by the 70+ age group whose monthly proportion of statin users increased from 22.3% - 35.6% between May 2009 and November 2021. During this period an increase in this age-group from 68.1% to 73.7% was seen for secondary prevention. In both cohorts, the monthly proportion of statin users was lowest among women, 25-39 age group, in those of black, mixed, and other ethnic groups, the most deprived group (primary prevention only) but highest in those with a history of myocardial infarction (secondary prevention only).

**Figure 2.**
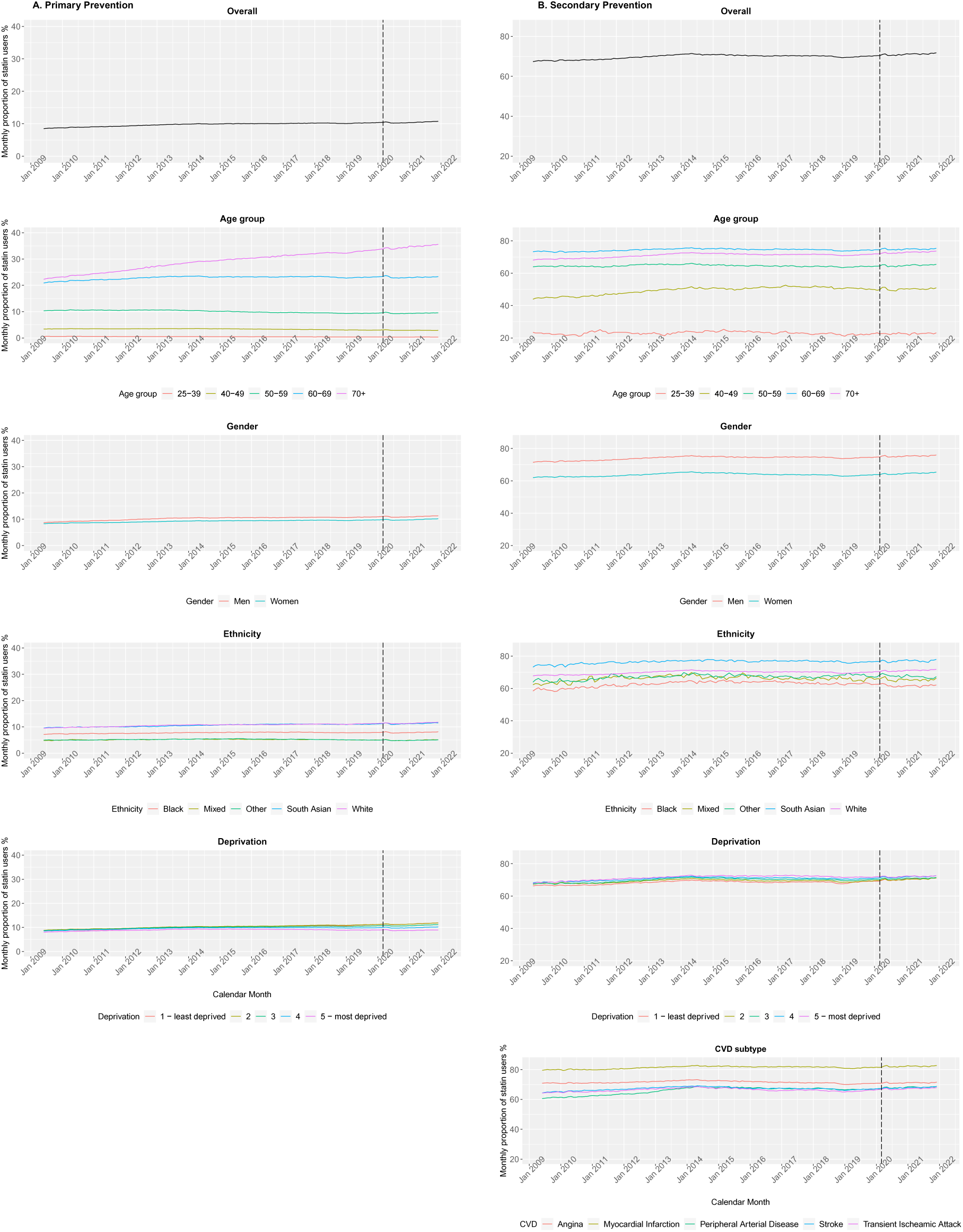
Monthly proportion of statin users for primary and secondary prevention Monthly proportion of statin users from May 2009 to November 2021 for primary and secondary prevention of cardiovascular disease. **A.** represents the primary prevention population and the denominator for this cohort are individuals without a history of cardiovascular disease. **B.** represents the secondary prevention population and the denominator for this cohort are individuals a cardiovascular disease diagnosis.

The monthly proportion of individuals with a GP recorded CVD risk assessment, regardless of eligibility for risk assessment, rose steadily from 13.7% in May 2009 to 35.3% in February 2020 and declined to 31.8% by November 2021 (Supplementary Figure 4). The proportion of CVD risk assessments, performed by GPs, was highest in the 40-74 age group and among those with less than 10% CVD risk score. Figure 3 shows that CVD risk assessment was most likely among the 70-74 age group compared to 40-49 age group adjusted HR 2.62 (95% CI, 2.60 − 2.65) and those of South Asian ethnicity adjusted HR 1.39 (95% CI, 1.38 − 1.40), Black ethnicity adjusted HR 1.28 (95% CI, 1.27-1.29) and Mixed ethnicity adjusted HR 1.15 (95% CI, 1.13-1.17) compared to white ethnicity. While there was no apparent trend across 4 lowest quintiles of deprivation, the most deprived group was more likely to receive a CVD risk assessment than the least deprived group adjusted HR 1.23 (95% CI, 1.22 − 1.24).

**Figure 3.**
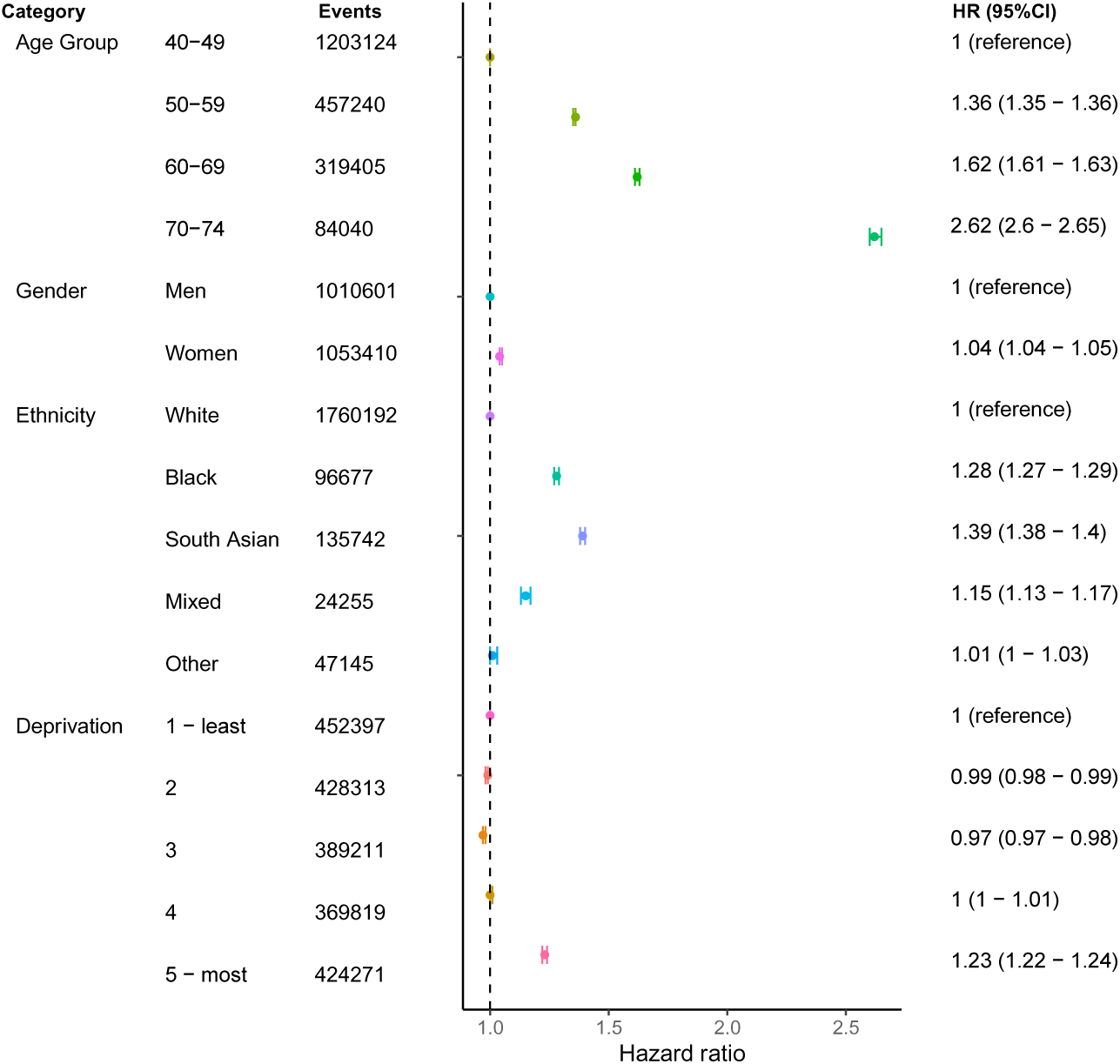
Adjusted hazard ratios for factors associated with receiving a recorded CVD risk assessment Hazard ratios from Cox proportional hazards regression model for the association between demographic factors and receiving a cardiovascular risk assessment, adjusted for gender, ethnicity and deprivation with age as the underlying time scale. Abbreviations: CI, confidence interval; HR, Hazard Ratio.

### Initiation

11.5% (n=49,877) of individuals with recorded risk score above the threshold initiated a statin within 28 days (primary prevention) while 79.0% (n=115,968) of individuals with a CVD diagnosis were initiated statins within 60 days of first CVD event (secondary prevention. Figure 4 shows that among individuals with an above threshold recorded CVD risk score, statin initiation was least likely in the 70+ age group than the 40-49 age group (adjusted OR 0.75, 95% CI, 0.72 − 0.79), people of black ethnicity than white ethnicity (adjusted OR 0.82, 95% CI: 0.77 − 0.88) but statin initiation was more likely in those in the most deprived group than in the least deprived group (adjusted OR 1.16, 95% CI; 1.12 − 1.20) and among women compared to men (adjusted OR 1.06, 95% CI; 1.04 – 1.08). In contrast, for secondary prevention, statin initiation within 60 days of a first CVD event was highest among the 60-69 age group than the 25-39 age group (adjusted OR 3.19, 95% CI; 3.02 − 3.38) and among people of south Asian ethnicity compared to those of white ethnicity (adjusted OR 1.53, 95% CI, 1.42-1.64). On the other hand, statin initiation was less likely among women compared to men (adjusted OR 0.70, 95% CI, 0.68 − 0.72) and in the black group compared to the white group (adjusted OR 0.71, 95% CI, 0.65-0.77). We also found that the factors associated with initiation were similar across CVD subtypes (Supplementary Figure 5).

**Figure 4:**
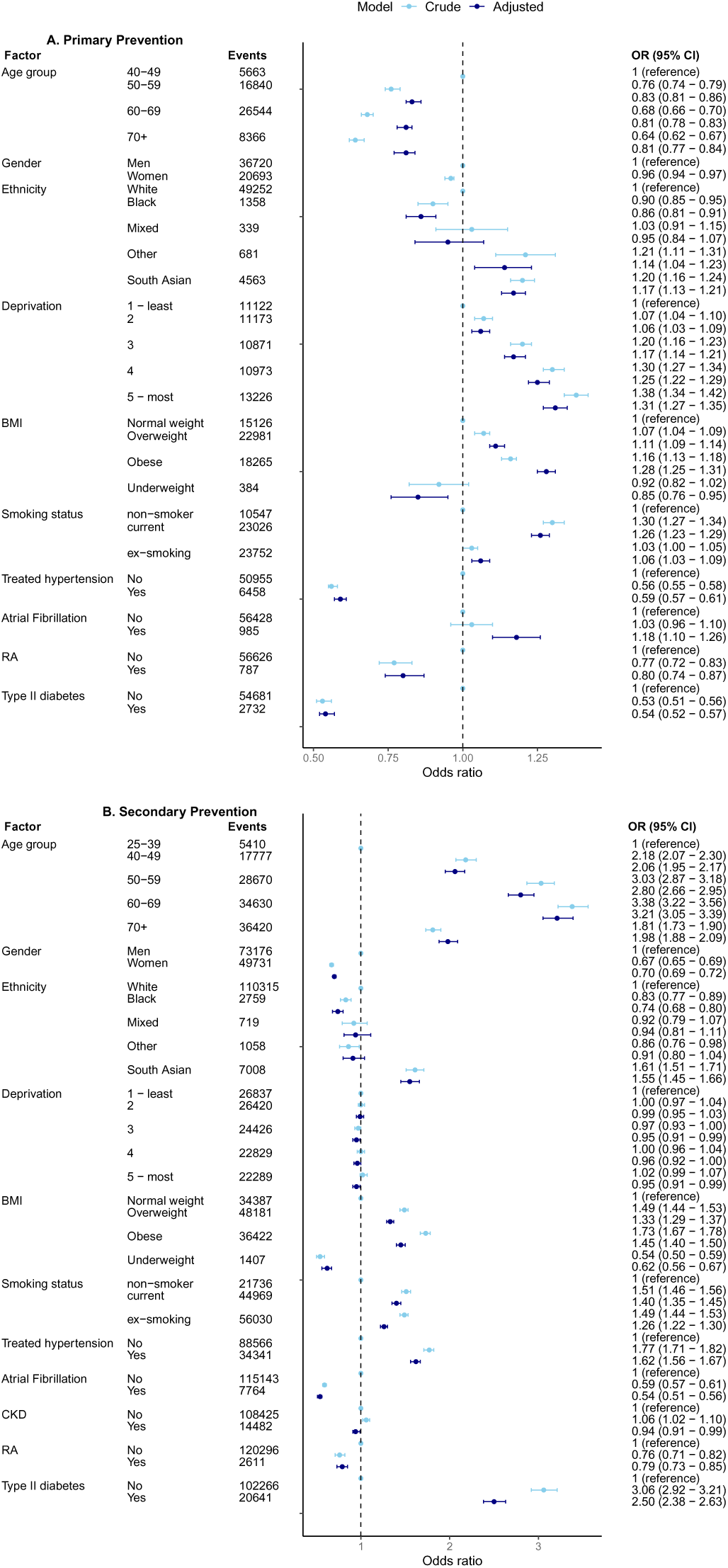
Odds ratios for the factors associated with statin initiation for primary and secondary prevention Odds ratios for factors associated with statin initiation for primary and secondary prevention. A. Primary prevention population which includes individuals with an above threshold GP recorded CVD risk score with statin initiation defined as having a statin prescription within 28 days of CVD risk score. B. Secondary prevention population comprising of individuals with a first CVD diagnosis during study period with statin initiation defined as having a statin prescription within 60 days of CVD diagnosis.

### Discontinuation

Among people who initiated a statin for primary prevention, 26.3% and 37.2% had discontinued statins by 1 year and 5 years of initiation, respectively. For secondary prevention, 10.0% and 19.9% had discontinued statins by 1 year and 5 years of initiation, respectively. We found that statin discontinuation was least likely in the 70+ year age group compared to the 25-39 age group (adjusted HR 0.65, 95% CI; 0.63 − 0.67) and slightly less likely among women compared to men (adjusted HR 0.97, 95% CI; 0.95 − 0.98) after adjusting for age, gender, ethnicity, and deprivation in the primary prevention cohort (Figure 5). Conversely, statin discontinuation was higher in the black group than the white group (adjusted HR 1.63, 95% CI; 1.59 − 1.68) and in the most deprived group compared with the least deprived group (adjusted HR 1.16, 95% CI; 1.14 − 1.18). In line with primary prevention findings, statin discontinuation among those with a history of CVD was most likely among people of black ethnicity compared to those of white ethnicity (adjusted HR 1.76, 95% CI; 1.65 − 1.89) and the most deprived group compared to the least deprived group (HR 1.08, 95% CI; 1.04 − 1.12) however women were also more likely to discontinue than men (HR 1.08, 95% CI; 1.06 − 1.11). Findings were also consistent with those according to CVD subtypes (Supplementary Figure 6). Kaplan Meier curves in Supplementary Figures 7 and 8 show that the time to statin discontinuation for primary and secondary prevention was shortest among the youngest age-groups, those of Black ethnicity, the most deprived socioeconomic groups, those with a CVD risk score below threshold (primary prevention only) and those with peripheral arterial disease (secondary prevention only). In addition, the curves were steepest for those with CVD risk score below threshold for initiation (primary prevention only) and for peripheral arterial disease (secondary prevention only).

**Figure 5.**
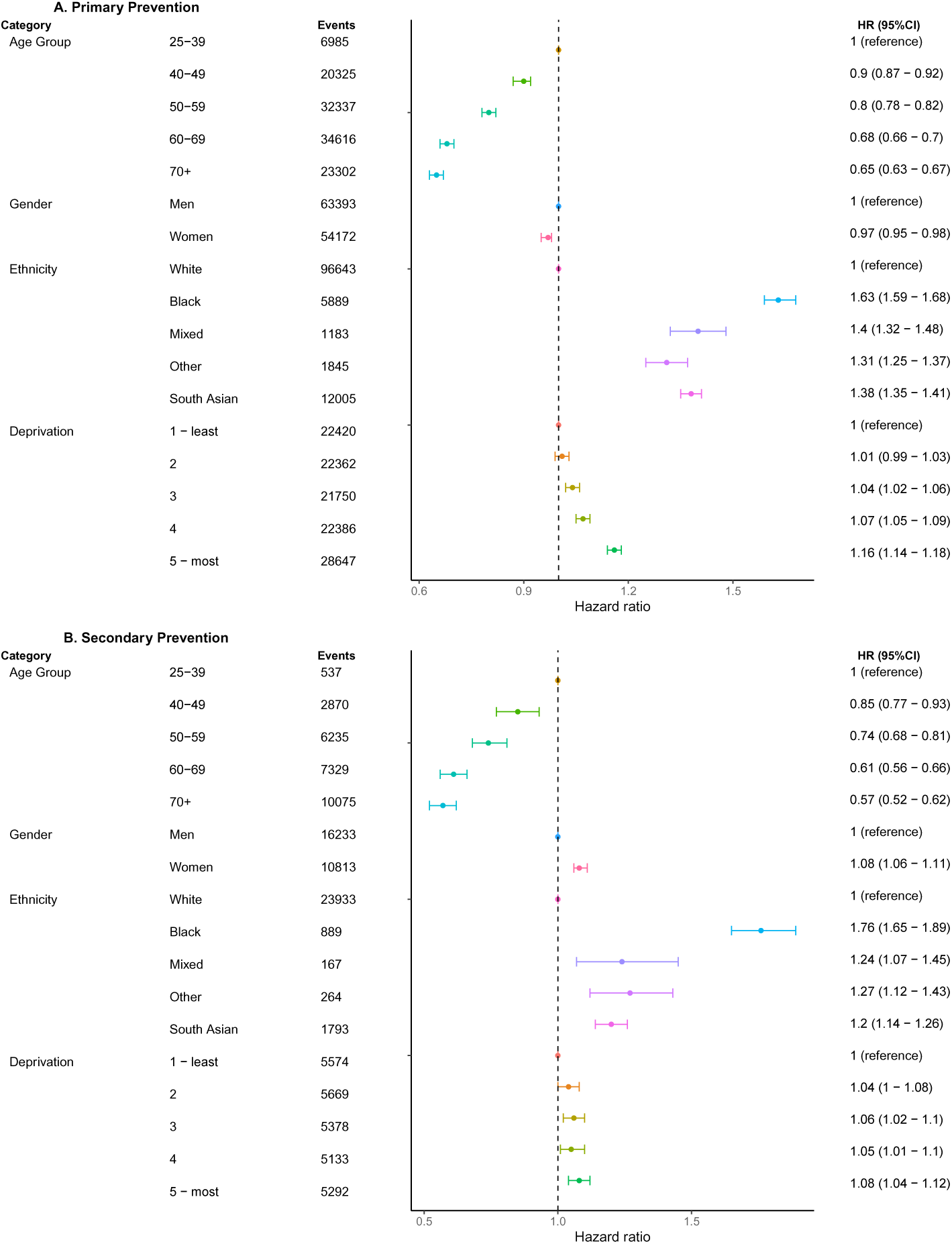
Adjusted Hazard Ratios for factors associated with statin discontinuation for primary and secondary CVD prevention. Hazard ratios from cox proportional hazards regression models for the association between demographic factors nd statin discontinuation (defined as reaching the end of a 90-day grace period without a new prescription) adjusted for gender, ethnicity and deprivation with age as the underlying time scale. **A.** represents the primary revention cohort which includes individuals aged 25 years and older who initiated statins without history of cardiovascular disease. Analyses are stratified by pre-pandemic period (before 1^st^ March 2020 and pandemic eriod (1^st^ March 2020 onwards). **B.** represents the cohort with a history of cardiovascular disease who initiated statins. Abbreviations: CI, confidence interval; HR, Hazard Ratio.

### Re-initiation

For both primary and secondary prevention, older people particularly those in the 70+ age group compared to 25-39 group, people of South Asian, black and mixed ethnicity compared to white people and people in the most deprived groups compared to the least deprived groups were most likely to restart statins with statin re-initiation highest in the 70+ age group compared to 25-39 age group adjusted HR 1.39 (95% CI, 1.25 – 1.55) for secondary prevention(Supplementary Figure 9). Women were less likely to re-initiate statins compared to men particularly for secondary prevention HR 0.87 (95% CI, 0.85 − 0.90). Similar associations were seen for all CVD subtypes except peripheral arterial disease (Supplementary Figure 10) and Kaplan-Meier curves depicting time from statin discontinuation to re-initiation are presented in Supplementary Figures 11 and 12).

### Number of CVD events prevented with optimal statin use and healthcare costs saved

Table 2 depicts the number of cardiovascular events that could be avoided based on current and 100% statin prevalence among those eligible for primary and secondary prevention. For primary prevention, 102,306 additional cardiovascular events would be prevented over 10 years if all eligible individuals were taking statins across England. For secondary prevention, 51,495 additional cardiovascular events could be prevented in England over 5 years if all individuals eligible were taking statins. If all CVD events from people currently not taking statins are avoided, £265.6 million, including the net cost of statins, can be saved for the NHS for primary prevention and £139.8 million can be saved for secondary prevention (Supplementary Table 6).

**Table 2.** Number of CVD events that can be prevented with optimal statin use.

**Primary Prevention based on 25% risk reduction<sup>a</sup>**
| GP recorded<br>Risk category | Number of<br>patients eligible | Current Statin<br>Prevalence | Total number of<br>expected events<br>based on risk score | Events avoided over<br>10 years based on<br>current prevalence | Events avoided over<br>10 years based on<br>100% prevalence | Additional events<br>avoided based on<br>100% prevalence | Scaled to<br>English<br>population |
| --- | --- | --- | --- | --- | --- | --- | --- |
| 10-19% | 384980 | 43% | 53660 | 13415 | 5828 | 7587 | 60726 |
| 20-29% | 144948 | 65% | 34928 | 8732 | 5719 | 3013 | 24113 |
| 30+% | 84656 | 73% | 32855 | 8214 | 6032 | 2182 | 17466 |
| Total | 614584 | 53% | 88588 | 30361 | 17578 | 12783 | 102306 |

**Secondary Prevention based on 10% absolute risk benefit over 5 years<sup>b</sup>**
| CVD subtype | Number of<br>patients eligible | Current Statin Prevalence | Events avoided over<br>5 years based on<br>current prevalence | Events avoided over 5<br>years based on 100%<br>prevalence | Additional events<br>avoided based on<br>100% prevalence | Scaled to<br>English<br>population |
| --- | --- | --- | --- | --- | --- | --- |
| MI | 73855 | 88% | 6464 | 7386 | 922 | 7379 |
| Angina | 71783 | 80% | 5734 | 7178 | 1445 | 11565 |
| TIA | 39052 | 77% | 3009 | 3905 | 896 | 7171 |
| Stroke | 44492 | 76% | 3402 | 4449 | 1047 | 8380 |
| PAD | 81278 | 74% | 6004 | 8128 | 2124 | 17000 |
| Total | 310460 | 79% | 24612 | 31046 | 6434 | 51495 |

**a.** risk reduction over 10 years for non-vascular disease derived from Collins R, Reith C, Emberson J, Armitage J, Baigent C, Blackwell L, et al. Interpretation of the evidence for the efficacy and safety of statin therapy. *Lancet*. 2016;388(10059):2532-61. **b.** 10% absolute risk benefit over 5 years derived from Armitage J, Baigent C, Barnes E, Betteridge DJ, Blackwell L, Blazing M, et al. Efficacy and safety of statin therapy in older people: a meta-analysis of individual participant data from 28 randomised controlled trials. *The Lancet*. 2019;393(10170):407-15. MI; Myocardial Infarction, TIA; Transient Ischaemic Attack, PAD; Peripheral Arterial Disease

### Additional secondary and sensitivity analyses

In additional secondary analyses, we stratified our main analyses by pre-pandemic and pandemic periods as shown in Supplementary Figures 13-16. These findings were broadly similar in the pre-pandemic and pandemic periods with larger confidence levels, owing to the smaller sample size, seen in the pandemic periods. Our sensitivity analyses were consistent with our main analyses as we observed minimal changes to our effect estimates (Supplementary Figures 17 and 18). Due to evidence of non-proportionality (Schoenfeld residuals test, p<0·05) for our Cox proportional hazards analyses on the association of demographic variables and risk of receiving a CVD risk assessment, statin discontinuation and statin initiation, we performed additional sensitivity analyses shown in Supplementary Tables 7-10. We found that risk of receiving a CVD was higher in the first 5 years from the start of follow up while individuals were less like to receive a CVD risk assessment more than 5 years after start of follow up. Statin discontinuation was less likely in the first 6 months of statin initiation in older age groups compared to more than 6 months and statin discontinuation was more likely among ethnic minority groups in the first 6 months than after 6 months for primary prevention with similar findings seen for secondary prevention. Analyses on re-initiation were consistent with our primary analysis.

## Discussion

In our comprehensive population-based cohort study of 5 million adults in England, we report the following key findings: 1) between May 2009 and November 2021, we saw an increase in statin prevalence for primary and secondary prevention among the 70+ age group while CVD risk assessment rose steadily over time from 15% to 32%, 2) the monthly prevalence of statins was lowest among black, mixed and other ethnicity and those in the most deprived socioeconomic quintile, however these groups, including people of south Asian ethnicity were more likely to receive a CVD risk assessment compared to the white ethnic group and least deprived groups, respectively, 3) 79.0% of individuals were initiated statins within 60 days of a CVD event. Statin initiation for secondary prevention was highest among people of South Asian ethnicity compared to those of white ethnicity but least likely among the black ethnic group compared to white ethnic group and among women compared to men, 4) 26.3% and 10.0% of individuals had discontinued statins within a year of statin initiation for primary and secondary prevention, respectively. Black people, the most deprived socioeconomic group and women (secondary prevention only) were most likely to discontinue statins compared to white people, least deprived groups, and men, respectively. These same groups including people of south Asian ethnicity and the 70+ age group compared to the white and 25-39 groups, respectively, were most likely to re-initiate statins for both primary and secondary prevention, 5) with optimal statin use among those eligible, around 100,000 additional cardiovascular events would be prevented over 10 years for primary prevention and 50,000 additional events over 5 years for secondary prevention. This would result in NHS healthcare cost savings of over £400 million.

### Comparison with other studies

Our prevalence estimates are in line with a UK primary care study which observed a slowed rate of increase in statin prescription rates from 2007 onwards.(34) Cardiovascular risk scoring increased over time in our study, consistent with previous studies, however we observed a decline in recorded risk scores during the pandemic period.(7, 35) The COVID-19 pandemic severely impacted primary care services, with a substantial reduction in primary care contacts for CVD after the introduction of population-wide restrictions.(36) In agreement with our findings, between 2019/20 and 2020/21, the proportion of people invited for an NHS Health Check declined by 82% with the proportion of people taking up an NHS Health Check invite also declining by 84%, leading to a backlog and suboptimal care. (37–39) Prior to the pandemic, findings from a systematic review showed inequalities in the NHS health check with coverage and uptake higher among older age groups and females between 2009 and 2016.(40) Similarly, we found that women and older age groups were more likely to receive a CVD risk assessment, which is part of the NHS health check. Inequalities in CVD risk assessment have implications for statin use as receiving a CVD risk assessment increases the likelihood of a statin prescription.(10)

We found that statin initiation was less likely in older age groups compared to the 40-49 age group, in our primary prevention cohort among people with a recorded CVD risk score. This finding is in conflict with an English study using The Health Improvement Network (THIN) primary care data which found that statin initiation rate increased with age up to 70-74 years old. (41) Unlike our study, the THIN study did not restrict the study population to those with a CVD risk score above threshold for statin initiation. Conditioning on statin eligibility threshold in our study may have resulted in other factors associated with statin initiation being more predictive of initiation than age which may explain the differences in findings. Among people with CVD, our findings were consistent with other studies which found that women and black people were less likely to initiate lipid-lowering drugs than men and white people, respectively, while the south Asian ethnic group is more likely to initiate statins compared to the white group.(42, 43) Previous studies have shown that statins are less likely to be prescribed in black ethnic groups compared to white ethnic groups. (44, 45) Similarly, our secondary prevention results of a lack of association between deprivation and statin initiation is consistent with another UK study.(42)

We found that a high proportion of those who discontinued statins re-initiated which is in agreement with a Scottish CPRD study that found that 72% and 75% of those who discontinued for primary and secondary prevention, respectively, also re-initiated statins. Consistent with our findings, the authors also reported that women and black, south Asian and other ethnic minority groups were more likely to discontinue statins compared to men and the white ethnic group, respectively.(46) For statin re-initiation, the study also found that women were less likely to re-initiate statins for secondary prevention than men while black and Asian groups were more likely to re-initiate statins than white group.(46)

Ethnic differences observed in our study may be a result of residual confounding as we were unable to account for factors such as cholesterol levels due to high level of missingness or understand the reasons for under-prescribing and discontinuation of statins. A previous UK study found that most patients initiated on statins had no previous CVD risk score. In the absence of a CVD risk score, cholesterol levels were found to be the main predictor of statin initiation.(7, 47) Prior studies have reported that people of black ethnicity are associated with lower than expected rates of access to and use of NHS cardiovascular care.(48) Evidence suggests that healthcare professionals experience considerable uncertainty in supporting ethnically diverse patients which leads to hesitance and contributes to ethnic health care inequalities.(49) Structural racism, discrimination and distrust have also been identified as key factors that negatively impact healthcare.(50) Deprivation has also been associated with CVD, with people living in the most deprived areas four times more likely to die prematurely from cardiovascular diseases.(51)

Statin discontinuation could have been affected by statin intensity as high-intensity statins could potentially increase the risk of side effects. However, a recent study in Scotland found that discontinuation was less likely in those treated with high intensity statins compared to moderate intensity.(52) Further studies are needed to explore this link in England. Additionally, concerns on the risks and benefits of statins, particularly following media coverage, may have contributed to an increase in statin discontinuation.(53)

### Strengths and Limitations

Our large cohort of 5 million individuals is the first to comprehensively assess inequalities across all stages of statin use in England for primary and secondary prevention to the best of our knowledge. Other strengths of our study include a long follow-up period of 13 years and the use of linked primary and secondary electronic records broadly representative of the UK population in terms of age, gender, ethnicity and geographical location and well recorded GP prescription data.(54)

There were several limitations in our study. Firstly, in our cardiovascular risk assessment, discontinuation, and re-initiation analyses, we did not adjust for non-fixed covariates such as BMI and smoking status due to a lack of time-updated information on these factors. Furthermore, data on cholesterol and blood pressure measurements had a high proportion of missing data thus these factors were not adjusted for in our initiation analyses. As a result, we cannot rule out residual confounding. Secondly, low intensity statins can be purchased over the counter and thus we were not able to capture information on statin use outside of primary care. Third, the reasons for statin discontinuation are not well captured in primary care records thus we were unable to explore this. Fourth, there is no data on whether individuals actually collected their statin prescription or took their medication thus misclassification of statin discontinuation is possible as we cannot be certain of when they discontinued statins.

Fifth, estimates on the number of cardiovascular events prevented and costs reduced rely on assumptions such as the cardiovascular risk and cost of hospitalisations remaining constant over time. Sixth, although we assessed changes in statin prescribing, we did not investigate whether there was a change in the incidence of cardiovascular events over time particularly during the COVID-19 pandemic though this has recently been reported.(36) We assessed this for primary prevention by observing changes in CVD risk category over the 13-year study period.

## Conclusion

We observed inequalities by age, gender, ethnicity and deprivation across multiple stages of statin use including statin prevalence, CVD risk scoring, initiation, discontinuation and re-initiation. Optimising statin use among those eligible, could lead to over 150,000 reduced CVD events leading to substantial NHS savings of over £400 million. Further research is needed to understand the reasons for under prescribing and discontinuation of statins particularly among women, black ethnic groups and the most deprived socioeconomic groups.

Our study suggests that there is an opportunity to optimise statin treatment through improving CVD risk assessment. Reductions in CVD risk assessment during the COVID-19 pandemic highlights the urgent need to improve uptake of routine CVD risk assessment to minimise missed opportunities for statin treatment thereby reducing preventable CVD events and additional costs to the NHS. This also has implications for pandemic planning to ensure important health services can be maintained during periods of reduced in person contact. Future qualitative studies are also needed to explore drivers of variation in statin adherence to understand which groups of patients are less likely to adhere to treatment and thus should be specifically targeted through education and prescribing strategies to reduce preventable CVD events in these groups.

## Data Availability

Data were obtained from the CPRD and are not publicly available. Access to CPRD data is dependent upon approval of a study protocol via CPRD RDG process (details at https://cprd.com/research-applications)

https://github.com/RutendoMuzambi/trends_statin_use

## Contributors

All authors contributed to the conception and/or design of this study. RM analyses the data and wrote the first draft of the manuscript. All authors were involved in the data interpretation and revision of the manuscript.

## Ethical approval

Our study protocol was approved via CPRD’s Research Data Governance (RDG) process (study ID 22_001872). Ethical approval was also obtained from the London School of Hygiene and Tropical Medicine research ethics committee (reference: 28002)

## Data sharing

Study protocol, analysis codes, codelists and additional results for this study can be accessed at https://github.com/RutendoMuzambi/trends_statin_use. Data were obtained from the CPRD and are not publicly available. Access to CPRD data is dependent upon approval of a study protocol via CPRD’s RDG process (details at https://cprd.com/research-applications)

## Transparency

The lead author affirms that this manuscript is an honest, accurate, and transparent account of the study being reported; that no important aspects of the study have been omitted; and that any discrepancies from the study as planned (and, if relevant, registered) have been explained.

