## Supplementary material for "Trends and inequalities in statin use for the primary and secondary prevention of cardiovascular disease between 2009 and 2021 in England"

### Supplementary Methods

**Data source and linkage**

We extracted data from the January 2022 CPRD (Clinical Practice Research Datalink) Aurum build consisting of over 40 million individuals as shown in Supplementary Figure 1. We extracted data from individuals eligible for linkage with the Hospital Episode Statistics (HES) Admitted Patient Care (APC) dataset and applied our exclusion criteria.

#### Supplementary Figure 1: Study flow diagram including linkage to HES APC dataset

Population in CPRD Aurum, Jan 2022 build **(N=** **40,833,738)**

<25 years old before end date (n=11,172,488)

*End date: Earliest of deathdate, last collection date, transfer out date, end of study observation period (31st Dec 2021*

25 years and older **(N=** **24,772,897)**

Less than 12 months research standard follow up between CPRD start date and end date (n=2,412,192)

Population with linked HES APC data **(N=** **35,945,385)**

Without linked HES data (N=4,888,353)

Male and female **(N=** **24,772,550)**

Sex other than male or female (n=347)

At least 12 months research standard follow up **(N=22,360,358)**

Follow up after 1/4/2009 **(N=** **16,650,277)**

Follow up in CPRD ends before 01/04/2009 (n= 5,710,081)

**Random subsample**

In total, there were 16,650,277 aged 25 years and older between 1^st^ April 2009 and 31^st^ December 2021 eligible for HES linkage with at least 12 months of research standard follow up in CPRD Aurum. We used STATA’s random-number generation function, runiform() which produces numbers that can pass for independent draws. We used this function to generate our subsample of 5 million individuals. The code for generating this subsample can be found on Github.(1)

#### Supplementary Table 1: Characteristics of individuals in original sample and random sample

|  | **Original Sample n (%)** | **Random subsample n (%)** |
| --- | --- | --- |
| **Total** | 16,650,277 | 5,000,000 |
| **Mean age (years)** | 44.46012 | 44.47283 |
| **Median age (years)** | 40 (29-56) | 40 (29-56) |
| **Sex** |  |  |
| Female | 50.4 | 50.4 |
| Male | 49.6 | 49.6 |
| **Region** |  |  |
| 1 | 2.83 | 2.84 |
| 2 | 16.93 | 16.93 |
| 3 | 3.34 | 3.35 |
| 4 | 1.96 | 1.95 |
| 5 | 15.07 | 15.05 |
| 6 | 4.12 | 4.13 |
| 7 | 24.18 | 24.18 |
| 8 | 19.93 | 19.91 |
| 9 | 11.65 | 11.66 |

**Statins data cleaning**

We extracted prescription data for statins specially simvastatin, atorvastatin, rosuvastatin, fluvastatin and	pravastatin. An individual was defined as a current statin user if they had an ongoing statin prescription based on the number of days prescribed, allowing for a grace period of 30 days. CPRD Aurum data contained a duration variable pertaining to how long medication was prescribed for. When the duration variable was zero, we calculated the duration of statin prescription using the quantity and daily dose. If the duration variable was zero and both the quantity and daily dose were not missing or coded as zero, duration was calculated by dividing the quantity by the daily dose. If the duration variable was zero but either the daily dose is zero or missing, and the quantity is not zero and not missing, then the duration was equal to the quantity. If duration is less than or equal to one, then the duration was 28 days. From these rules we created a variable for when statin prescriptions would be due to run out. If there were multiple statin prescriptions on same date for the same patient, the prescription with the longest duration for a given date was kept. If the duration of the statin prescription was coded as more than 180 days, then the duration was	coded as the median duration which was 28 days.

**Missing data**

The table below summaries the covariates with missing values and the proportion of missingness for these covariates.

#### Supplementary Table 2: Proportion of missing covariates

| **Proportion of missing covariates in study** | |
| --- | --- |
| **Covariate** | **Missing N (%)** |
| Ethnicity | 371,583 (7.4%) |
| Deprivation | 7,183 (0.1%) |
| BMI | 626,445 (12.5%) |
| Smoking Status | 161,291 (3.2%) |
| Systolic Blood Pressure (SBP) | 780,089 (15.6%) |
| Diastolic Blood Pressure (DBL) | 780,089 (15.6%) |
| High Density Lipoprotein (HDL) | 3,297,133 (65.9%) |
| Low Density Lipoprotein (LDL) | 3,562,367 (71.2%) |
| Total Cholesterol | 3,086,709 (61.7%) |
| Total Cholesterol: HDL ratio | 3,599,755 (72.0%) |

These data above are unlikely to be missing at random as GPs are more likely to record information with clinical implications thus multiple imputation may be biased. We therefore conducted a complete case analysis for our initiation analyses which is valid providing missingness is independent of the outcome under study. Missingness for blood pressure and cholesterol measurements was too high (>20%) for the covariates to be included adjusted for in the initiation analyses.

**NHS Reference Costs**

To calculate costs of cardiovascular events, we used the national schedule of NHS costs financial year 2021-2022. We specifically calculated the costs of myocardial infarction, stroke, angina and transient ischaemic attack whose costs from all NHS trusts and NHS foundation trusts were included in the total healthcare resource groups. The calculated the total costs of all CC (comorbidity and complication) scores for event CVD event using the total cost columns provided.

| Supplementary Table 3: National Schedule of NHS costs financial year 2021-22 | | | | |
| --- | --- | --- | --- | --- |
| **Currency** | **Currency description** | **Activity** | **Unit cost** | **Total cost** |
| AA35A | Stroke with CC Score 16+ | 40,247 | £7,319 | £294,569,844 |
| AA35B | Stroke with CC Score 13-15 | 29,669 | £5,246 | £155,634,064 |
| AA35C | Stroke with CC Score 10-12 | 31,026 | £3,920 | £121,622,177 |
| AA35D | Stroke with CC Score 7-9 | 28,829 | £2,960 | £85,341,544 |
| AA35E | Stroke with CC Score 4-6 | 23,495 | £2,335 | £54,851,198 |
| AA35F | Stroke with CC Score 0-3 | 13,820 | £1,780 | £24,593,554 |
| EB10A | Actual or Suspected Myocardial Infarction, with CC Score 13+ | 23,437 | £3,195 | £74,871,331 |
| EB10B | Actual or Suspected Myocardial Infarction, with CC Score 10-12 | 23,710 | £2,333 | £55,307,577 |
| EB10C | Actual or Suspected Myocardial Infarction, with CC Score 7-9 | 25,991 | £1,877 | £48,776,366 |
| EB10D | Actual or Suspected Myocardial Infarction, with CC Score 4-6 | 27,531 | £1,655 | £45,566,919 |
| EB10E | Actual or Suspected Myocardial Infarction, with CC Score 0-3 | 21,118 | £1,386 | £29,271,801 |
| EB13A | Angina with CC Score 12+ | 8,445 | £1,582 | £13,359,738 |
| EB13B | Angina with CC Score 8-11 | 14,528 | £1,177 | £17,100,653 |
| EB13C | Angina with CC Score 4-7 | 18,657 | £919 | £17,148,112 |
| EB13D | Angina with CC Score 0-3 | 9,949 | £708 | £7,039,615 |
| AA29C | Transient Ischaemic Attack with CC Score 11+ | 7,718 | £2,176 | £16,792,659 |
| AA29D | Transient Ischaemic Attack with CC Score 8-10 | 5,092 | £1,293 | £6,585,412 |
| AA29E | Transient Ischaemic Attack with CC Score 5-7 | 6,232 | £1,035 | £6,450,249 |
| AA29F | Transient Ischaemic Attack with CC Score 0-4 | 7,667 | £781 | £5,990,240 |

**Prescription Cost Analysis**

The Prescription Cost Analysis contains details of costs and volumes of prescriptions dispensed in the community in England. The table below contains the information obtained to calculate costs of first-line statins for primary (atorvastatin 20mg) and secondary prevention (atorvastatin) according to the British National Formulary (BNF) for preparation class 01.

#### Supplementary Table 4: Prescription cost analysis for Atorvastatin 20mg and Atorvastatin 80mg tablets

| **BNF Presentation Code** | **BNF Presentation Name** | **Total Items** | **Total Quantity** | **Total Cost (GBP)** | **Cost Per Item (GBP)** | **Cost Per Quantity (GBP)** | **Quantity Per Item** |
| --- | --- | --- | --- | --- | --- | --- | --- |
| 0212000B0AAABAB | Atorvastatin 20mg tablets | 24,835,499 | 835,620,526 | 35,081,171.26 | 1.41 | 0.04 | 33.65 |
| 0212000B0AAADAD | Atorvastatin 80mg tablets | 7,147,599 | 204,711,835 | 12,809,578.56 | 1.79 | 0.06 | 28.64 |

### Supplementary Results: Figures and Tables

#### Supplementary Figure 2: Flowchart of study populations for primary prevention

Total population aged 25 years and older with CPRD Aurum data linked to HES APC with at least 12 months research standard follow up between 1st April 2009 and 31 December 2021 (N=16,650, 277)

Those not included in random subsample (N=11,650,277)

Without history of CVD or statin contraindications before start of follow up (N=4,658,400)

Alive in the study on or after 30/04/2009 (N=4,597,500)

Prevalence (Individuals without a history of CVD or statin contraindications)

Without a history of CVD diagnosis, CKD, T1DM, familial hypercholesterolaemia or statin contraindications at start of follow up and aged 25-84 years old. (N=4,470,870) Monthly proportion with CVD risk assessment cohort

Aged 40-74 before start of follow up (N=2,241,159) Factors associated with receiving CVD risk assessment cohort

With a recorded CVD risk score above threshold for statin initiation and on or after start of follow up aged 40-74 at CVD risk assessment (N=537,219)

Without exclusion event before recorded risk score and with risk score who remain alive during 28-day grace period and without a statin prescription before risk date (N=432,686)

Initiation (among those with eligible for primary prevention with recorded CVD risk above threshold)

With a first ever statin prescription after start of follow up & during study period (N=373,266)

Without CVD prior to or on the same day as statin initiation (N=303,292)

Discontinuation (among those who initated statins regardless of risk assessment)

Initiated statins during study period with no history of CVD (N=303,292)

Discontinued during study period, Initiated statins for without history of CVD, or CVD event prior to discontinuation(N=116,468)

Re-initiation (among those who initiated for primary prevention and discontinued)

Supplementary Figure 3: Flowchart of study populations for secondary prevention

Total population aged 25 years and older with CPRD Aurum data linked to HES APC with at least 12 months research standard follow up between 1st April 2009 and 31 December 2021 (N=16,650, 277)

Those not included in random subsample (N=11,650,277)

With a history of CVD before or during study period and without statin contraindications (N=425,112)

Statins issued on or after CVD event or not prescribed statins and alive after 30/04/2009 (N=406,746)

Prevalence (Individuals with a CVD event during the study period)

With a CVD event on or after start of follow up and before end of study (N=156,100)

Remain alive during 60-day grace period after CVD event (N=146,811)

Initiation (among those with a CVD event during study period and alive 60-days after CVD event)

With a CVD event on or after start of follow up and before end of study (N=165,953)

Initiated on statins during study period and after CVD event (N=122,743)

Discontinuation (among those with CVD event and statin prescription after event)

Initiated statins and had history of CVD event during study period (N=122,743)

Discontinued statins (N=27,257)

Re-initiation (among those with CVD event who discontinued statins)

#### Supplementary Table 5: Characteristics of individuals included in CVD risk assessment cohorts

| **Characteristics** | **Monthly prevalence of individuals with a CVD risk assessment** | **Factors associated with CVD risk assessment** |
| --- | --- | --- |
| **Total (N)** | 4,470,870 | 2,241,159 |
| **Median Age (years, IQR)** | 38.0 (29.0-52.0) | 47.0 (40.0 to 57.0) |
| **Mean Age (years)** | 41.95 14.97 | 49.0 (11.3) |
| **Age Group (years)** |  |  |
| 25-39 | 2,442,680 (54.6%) | 0 (0) |
| 40-49 | 822,846 (18.4%) | 1321138 (58.9) |
| 50-59 | 568,580 (12.7%) | 494799 (22.1) |
| 60-69 | 389,164 (8.7%) | 337440 (15.1) |
| 70-74 | 196,461 (4.4%) | 87782 (3.9) |
| **Gender** |  |  |
| Men | 2,222,936 (49.7%) | 1106176 (49.4) |
| Women | 2,247,934 (50.3%) | 1134983 (50.6) |
| **Ethnicity** |  |  |
| White | 3,337,120 (74.6%) | 1762505 (85.3) |
| South Asian | 353,611 (7.9%) | 135826 (6.6) |
| Black | 210,014 (4.7%) | 96776 (4.7) |
| Mixed | 69,245 (1.5%) | 24278 (1.2) |
| Other | 143,571 (3.2%) | 47200 (2.3) |
| **Deprivation** |  |  |
| 1. Least deprived | 792,016 (17.7%) | 491662 (22.0) |
| 2 | 789,869 (17.7%) | 463117 (20.7) |
| 3 | 806,740 (18.0%) | 422559 (18.9) |
| 4 | 881,226 (19.7%) | 402024 (18.0) |
| 5. Most Deprived | 1,194,798 (26.7%) | 458805 (20.5) |
| **Baseline BMI, kg/m2** |  |  |
| Underweight (<18·5) | 98,260 (2.2%) | 30288 (1.4) |
| Normal weight (18·5–24·9) | 1,741,249 (38.9%) | 791483 (35.3) |
| Overweight (25·0–29·9) | 1,253,325 (28.0%) | 731957 (32.7) |
| Obese or morbidly obese (≥30·0) | 793,935 (17.8%) | 482186 (21.5) |
| Missing | 584,101 (13.1%) | 205245 (9.2) |
| **Smoking Status** |  |  |
| Non-smoker | 1,376,133 (30.8%) | 683878 (30.5) |
| Current smoker | 1,391,267 (31.1%) | 693504 (30.9) |
| Former smoker | 153,503 (3.4%) | 801444 (35.8) |
| Missing | 1,549,967 (34.7%) | 62333 (2.8) |
| **Systolic blood pressure** | 125.2 (15.7) | 127.9 (15.8) |
| **Diastolic blood pressure** | 76.3 (9.9) | 78.1 (9.7) |
| **HDL** | 1.4 (0.4) | 1.4 (0.4) |
| **LDL** | 3.1 (1.0) | 3.1 (0.9) |
| **TCHL** | 5.1 (1.1) | 5.2 (1.1) |
| **THDL** | 3.9 (1.3) | 3.9 (1.3) |
| **Hypertension** | 494,094 (11.1%) | 337031 (15.0) |
| **Treated Hypertension** | 300,543 (6.7%) | 204226 (9.1) |
| **Type II diabetes mellitus** | 157,074 (3.5%) | 100137 (4.5) |
| **Atrial Fibrillation** | 38,475 (0.9%) | 20813 (0.9) |
| **Rheumatoid Arthritis** | 32,386 (0.7%) | 21067 (0.9) |

#### Supplementary Figure 4: Monthly proportion of individuals who received a CVD risk assessment in the last 5 years

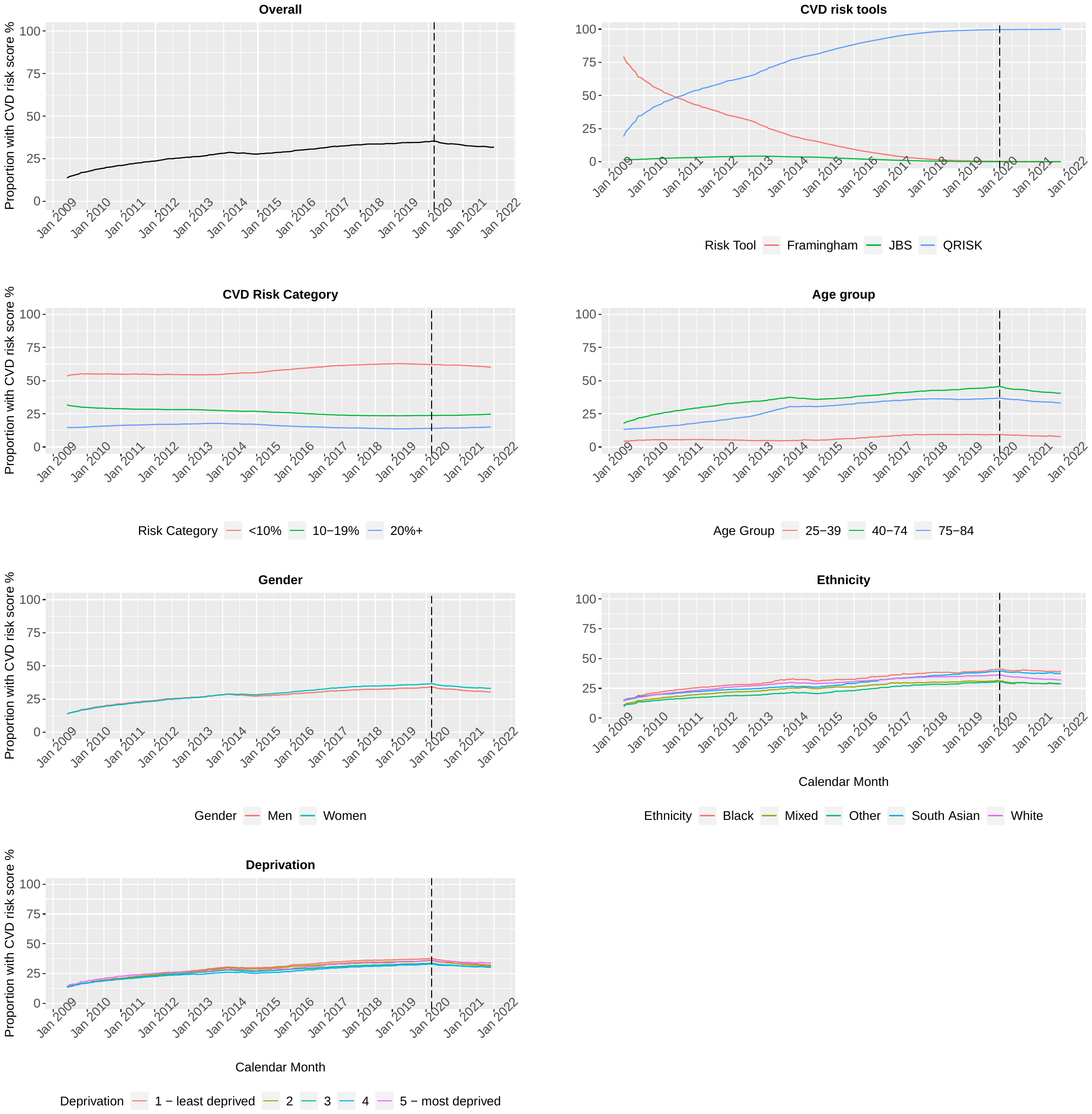

Monthly proportion of individuals with a current CVD (cardiovascular disease) risk record i.e. a CVD risk assessment in the last five years from May 2009 to November 2021 for primary and secondary prevention of CVD. Denominator includes all individuals aged with no history of CVD, chronic kidney disease, type 1 diabetes mellitus, familial hypercholesterolaemia and no contraindications to statins prior to the start of follow up.

#### Supplementary Figure 5: Odds ratios for the factors associated with statin initiation stratified by CVD subtype

Odds ratios (OR) for factors associated with statin initiation for secondary prevention stratified by cardiovascular disease subtype i.e myocardial infarction (MI), stroke, angina, TIA (transient ischaemic attack) and PAD (peripheral arterial disease). ORs presented were adjusted for age, gender, ethnicity, deprivation, BMI, smoking status., treated hypertension (defined as a hypertension diagnosis with subsequent antihypertive prescription), atrial fibrillation, chronic kidney disease (CKD), rheumatoid arthritis (RA) and type II diabetes mellitus.

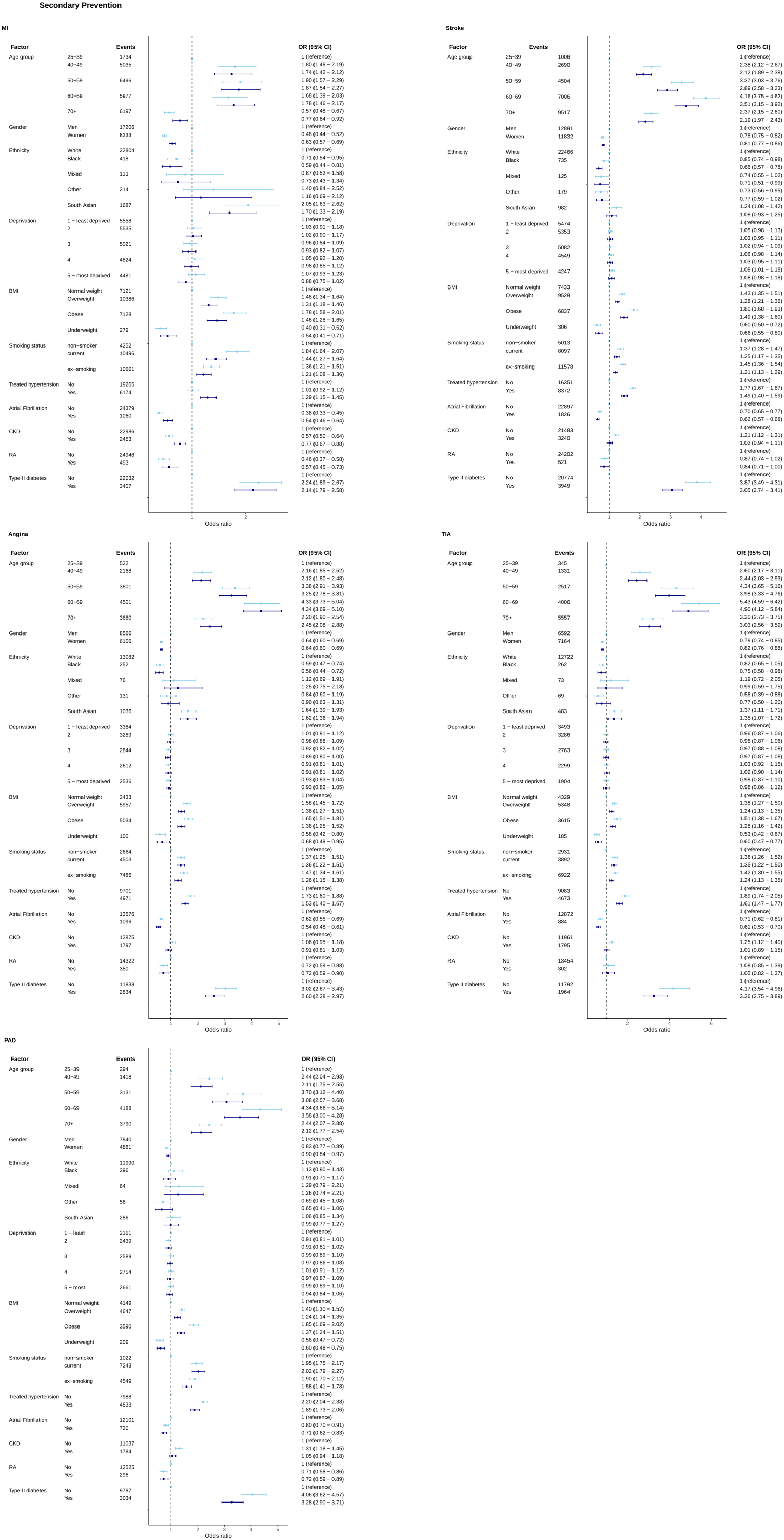

#### Supplementary Figure 6: Adjusted Hazard Ratios for factors associated with statin discontinuation stratified by CVD subtype

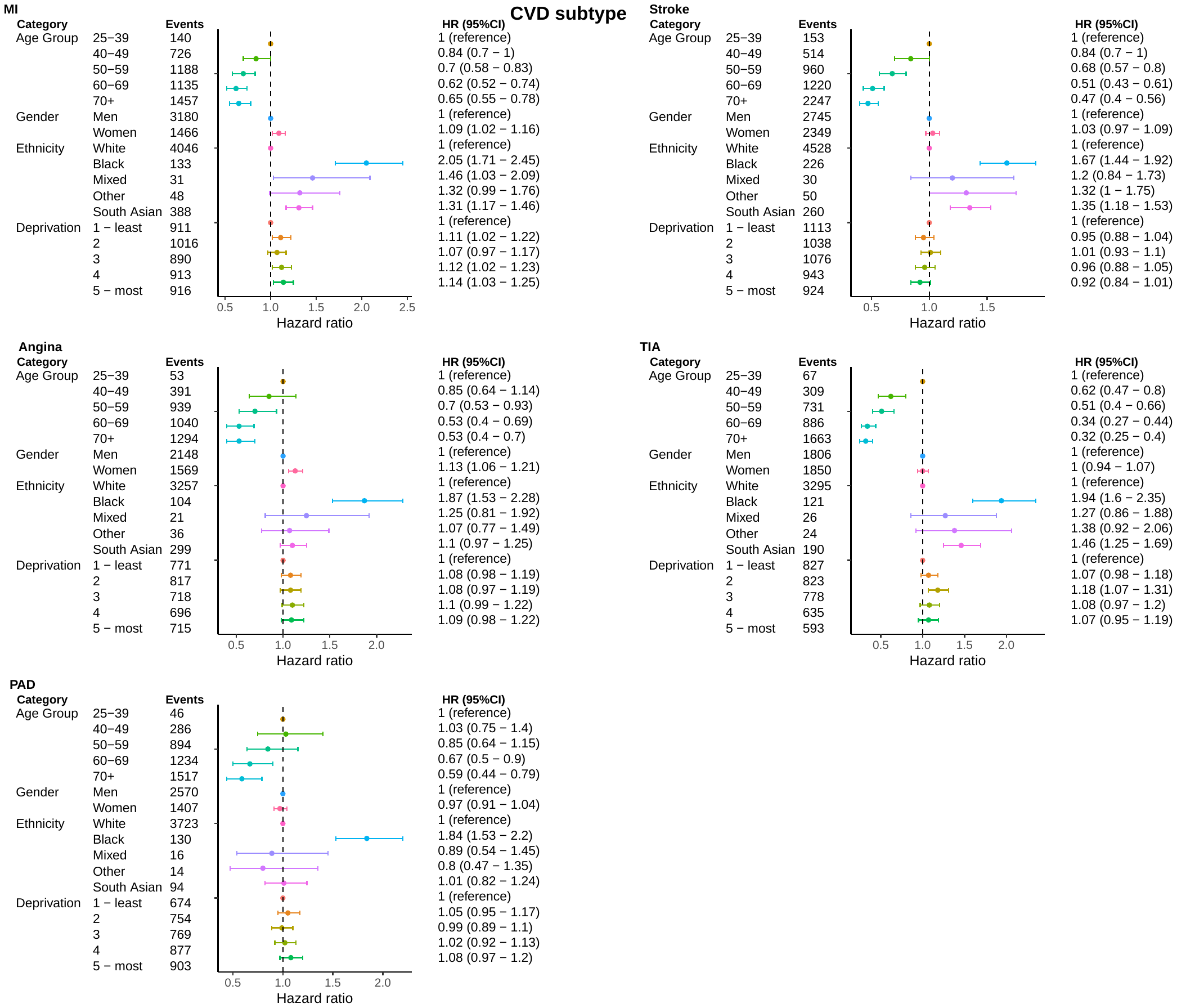

Hazard ratios (HR) from Cox proportional hazards regression models for the association between demographic factors and statin discontinuation for secondary prevention stratified by cardiovascular disease subtype i.e myocardial infarction (MI), stroke, angina, TIA (transient ischaemic attack) and PAD (peripheral arterial disease). Analyses were adjusted for age at statin initiation, gender, ethnicity and deprivation. Study population includes individuals with a CVD diagnosis during the study period who initiated statins and remained alive and under follow-up within 60 days of a CVD event.

#### Supplementary Figure 7: Kaplan Meier curves depicting the time to statin discontinuation for primary prevention

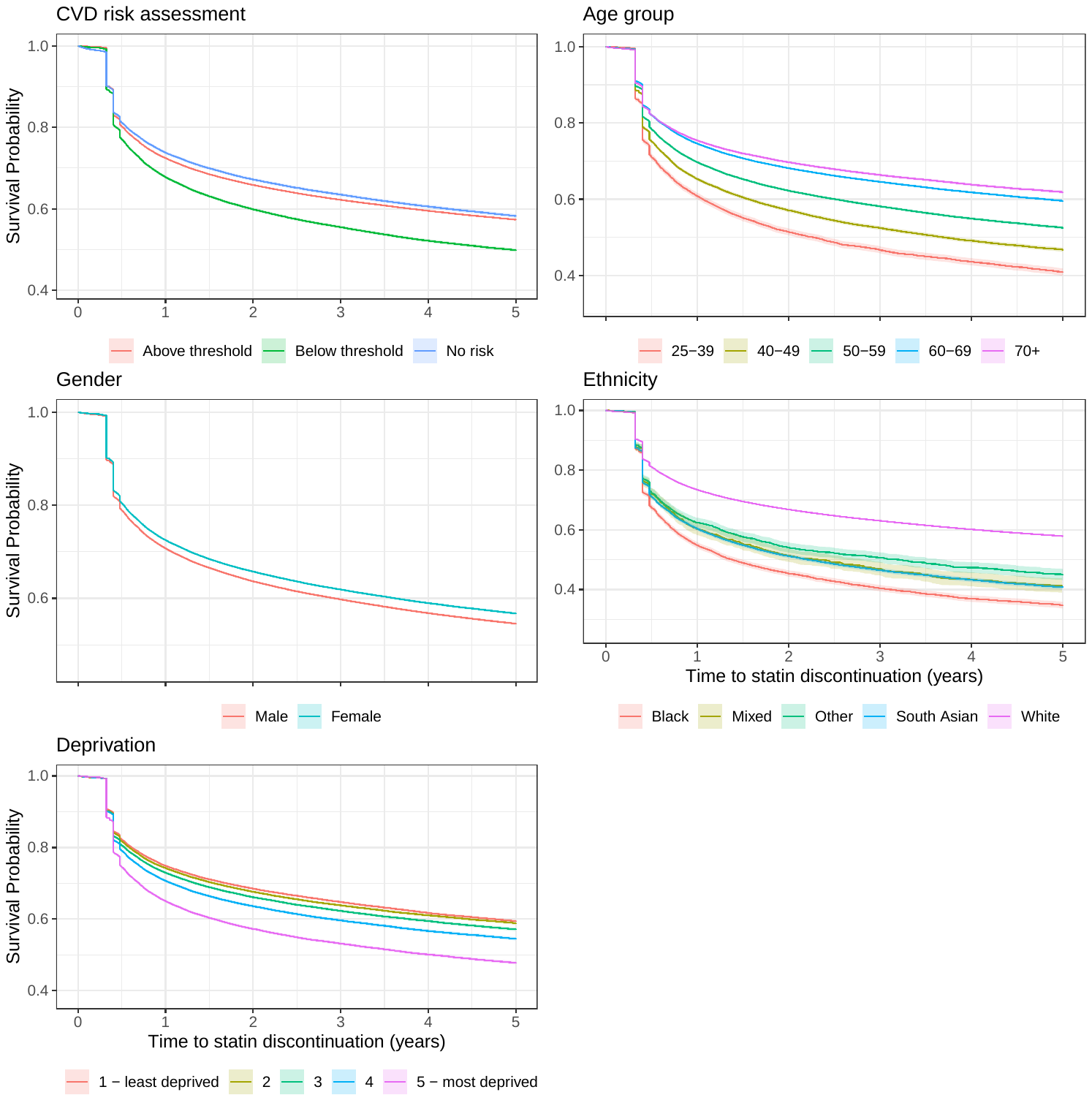

Kaplan Meier curves depicting time to statin discontinuation (years) among those who initiated statins for primary prevention i.e. without a pre-existing CVD diagnosis and with either no CVD risk assessment, above threshold (20 or greater pre 2014 and 10% or higher from 2014 onwards) and below threshold.

#### Supplementary Figure 8: Kaplan Meier curves depicting the time to statin discontinuation for secondary prevention

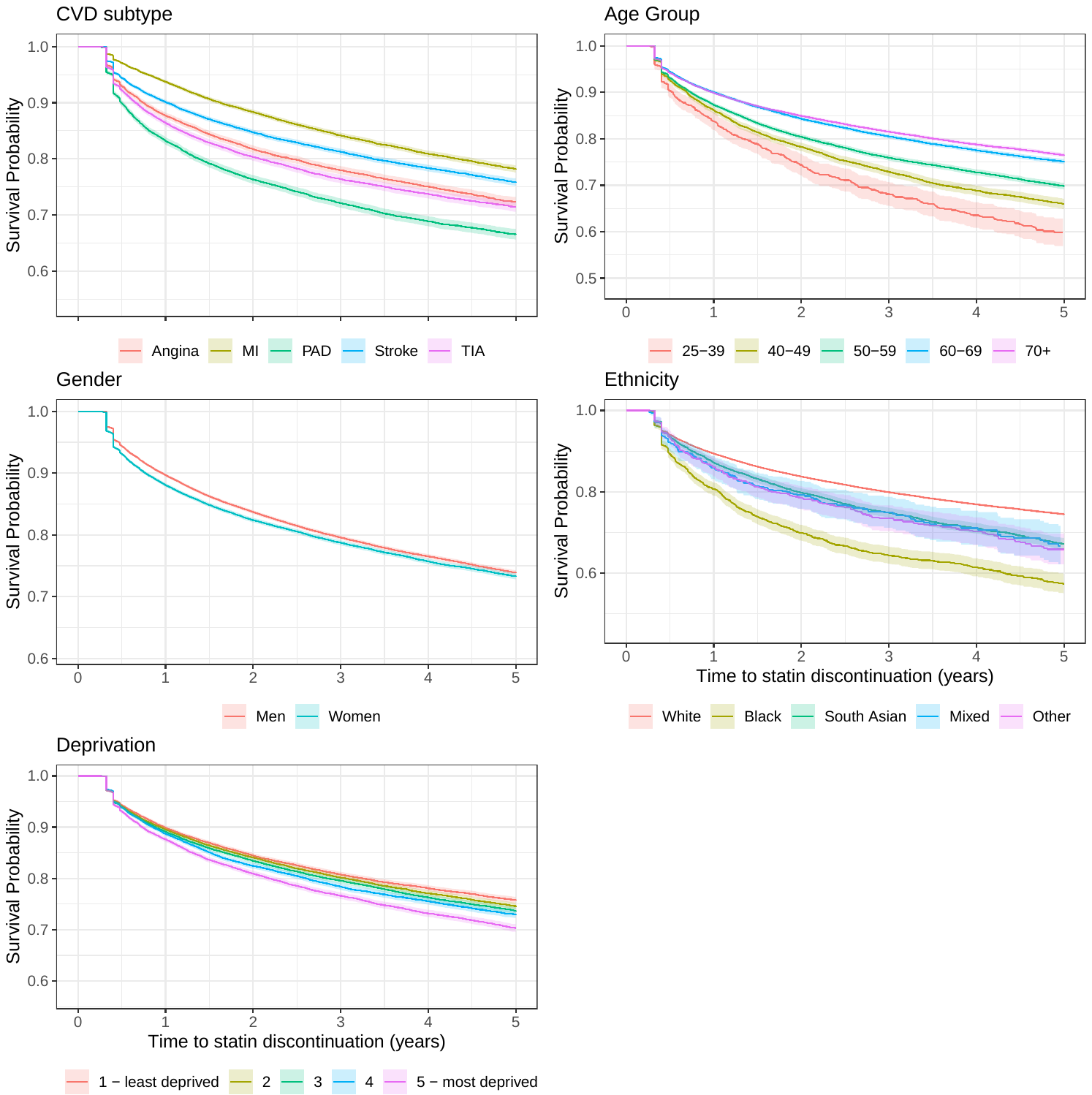

Kaplan Meier curves depicting time to statin discontinuation (years) among those who initiated statins for secondary prevention i.e. with a CVD diagnosis during the study period. Discontinuation defined as a reaching the end of a 90-day grace period without a new statin prescription.

#### Supplementary Figure 9: Adjusted Hazard Ratios for factors associated with statin re-initiation for primary and secondary CVD prevention

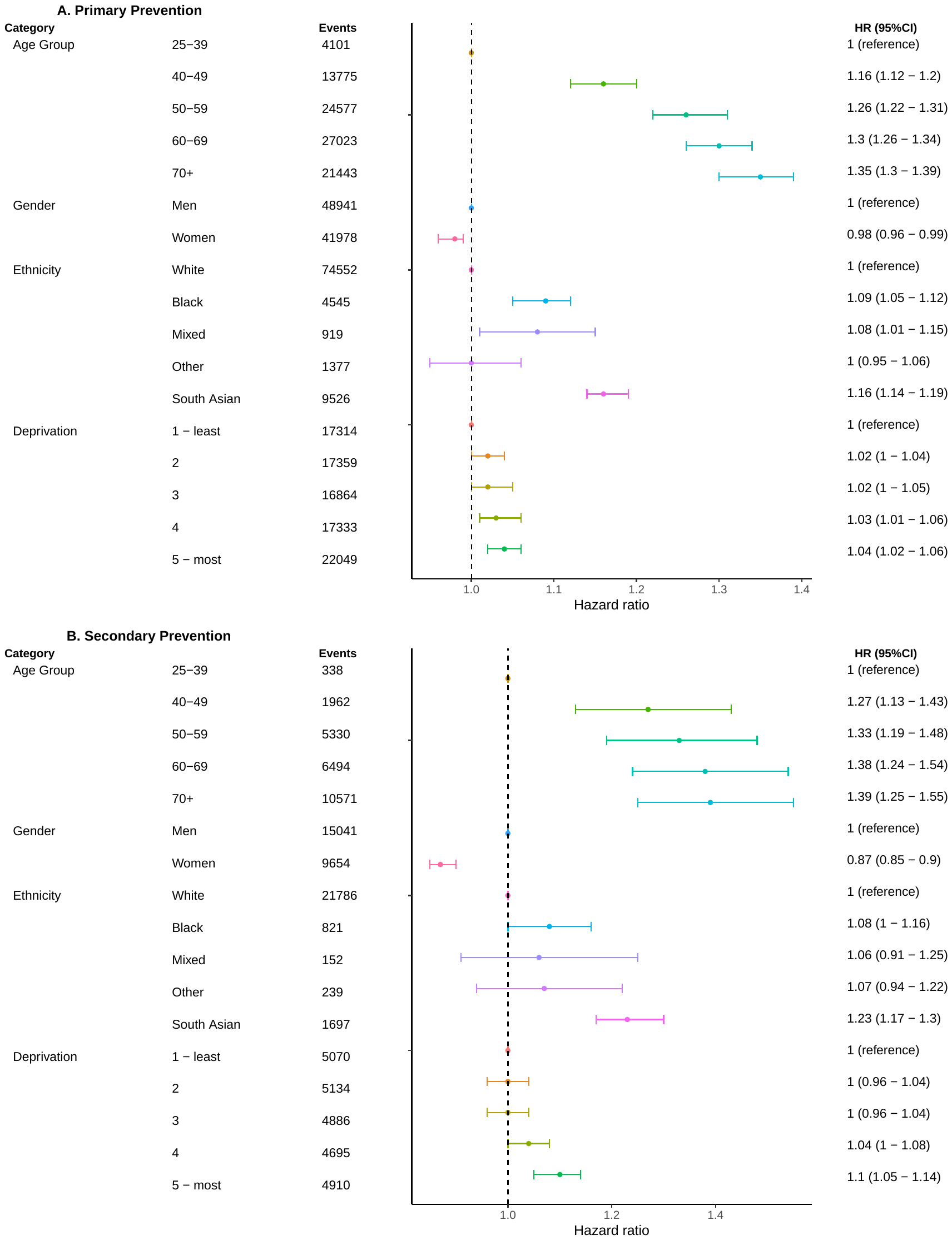

Hazard ratios (HR) from Cox proportional hazards regression models for the association between demographic factors and statin re-initiation for primary and secondary prevention. **A.** Primary prevention cohort with an existing CVD diagnosis prior to statin initiation and prior to statin cessation. **B.** Secondary prevention cohort with a CVD diagnosis during study period who initiated and discontinued statins. Re-initiation defined as a subsequent statin prescription after a 90 day period without a statin prescription. Analyses were adjusted for age at statin discontinuation, gender, ethnicity and deprivation.

#### Supplementary Figure 10: Adjusted Hazard Ratios for factors associated with statin re-initiation stratified by CVD subtype

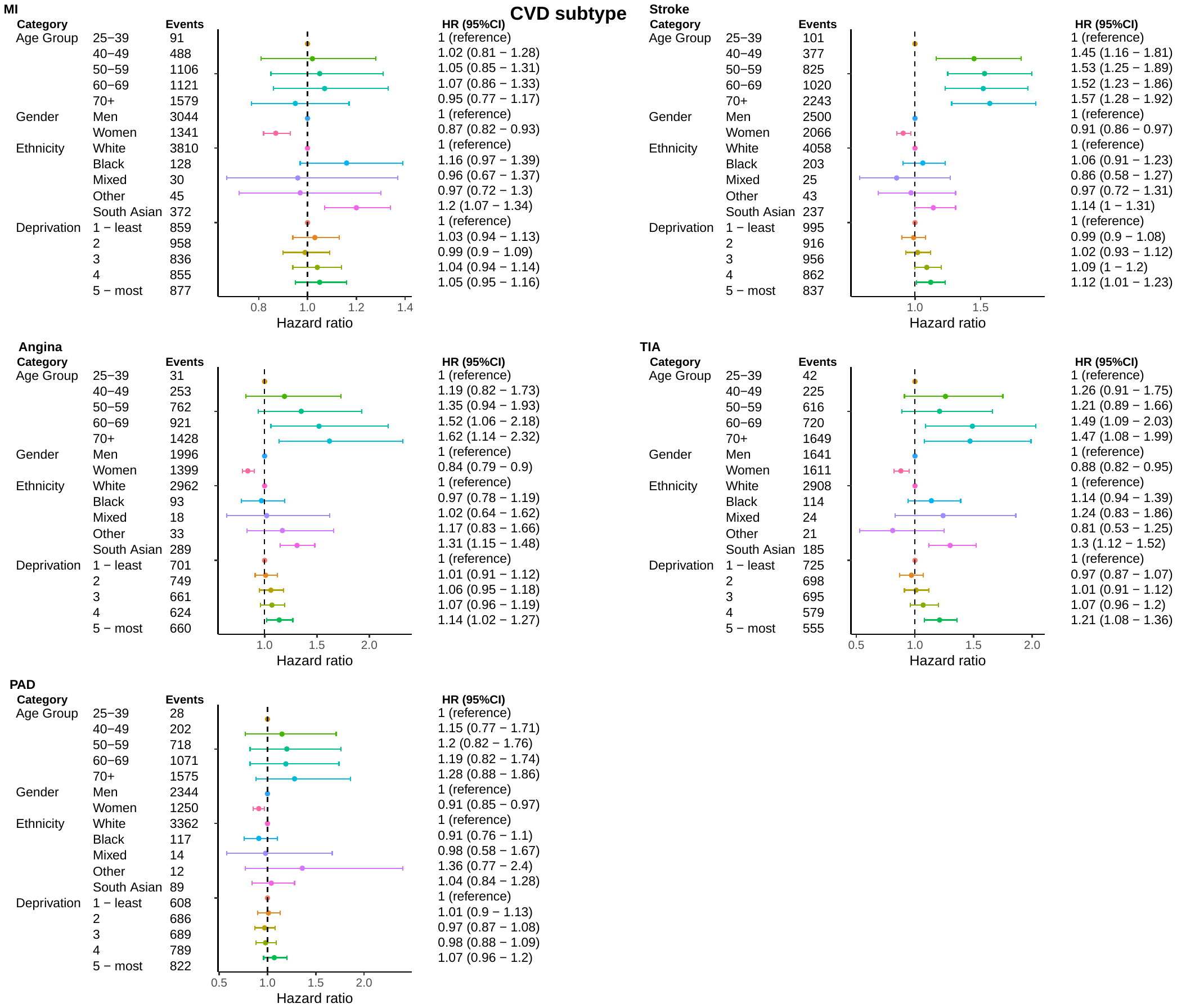

Hazard ratios (HR) from Cox proportional hazards regression models for the association between demographic factors and statin re-initiation for secondary prevention stratified by cardiovascular disease subtype i.e myocardial infarction (MI), stroke, angina, TIA (transient ischaemic attack) and PAD (peripheral arterial disease). Analyses were adjusted for age at statin initiation, gender, ethnicity and deprivation. Study population includes individuals with a CVD diagnosis during the study period who initiated statins and remained alive and under follow-up within 60 days of a CVD event.

#### Supplementary Figure 11: Kaplan Meier curves depicting the time to statin re-initiation taking statins for primary prevention

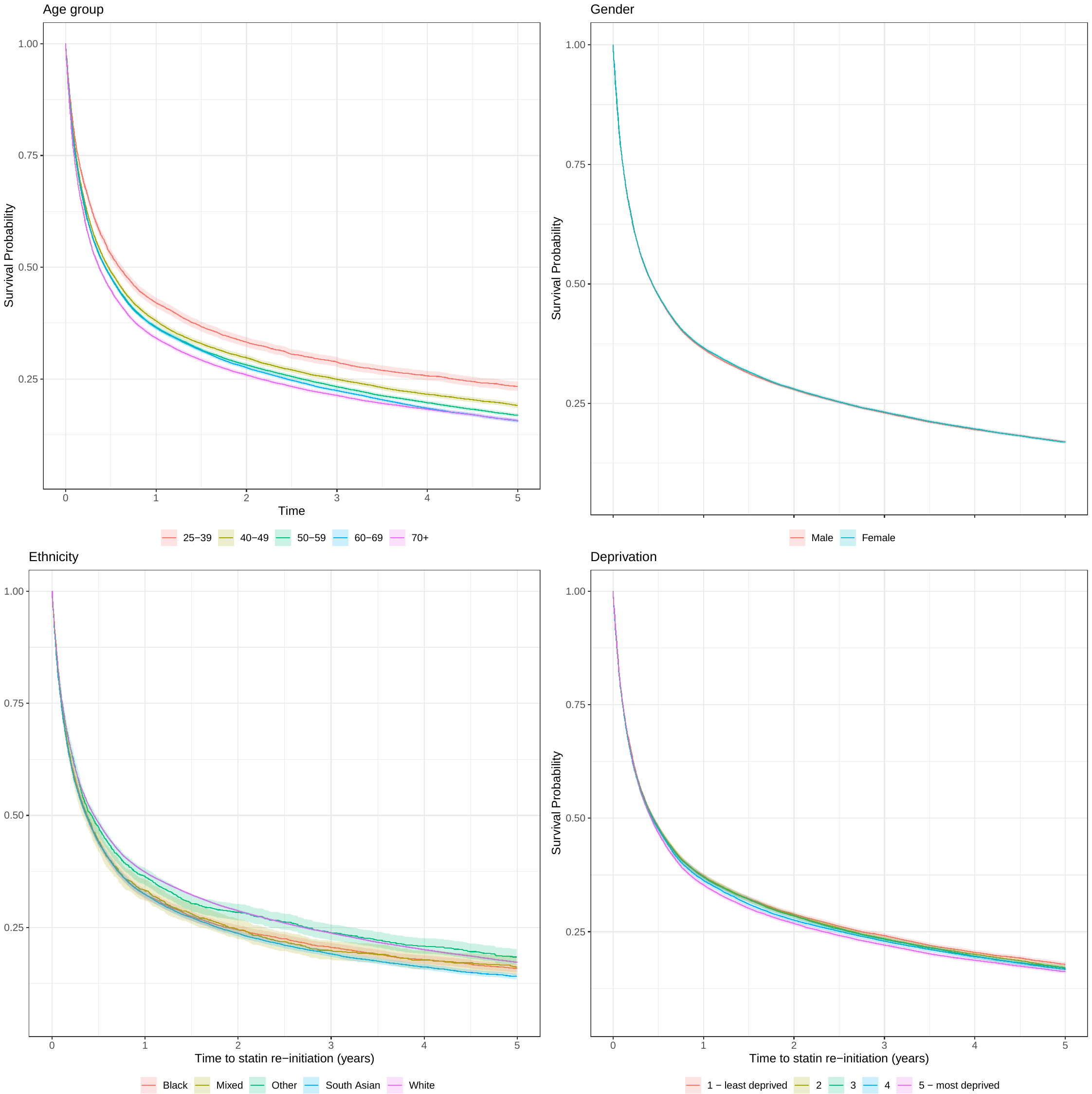

#### Supplementary Figure 12: Kaplan Meier curves depicting the time to statin re-initiation taking statins for secondary prevention

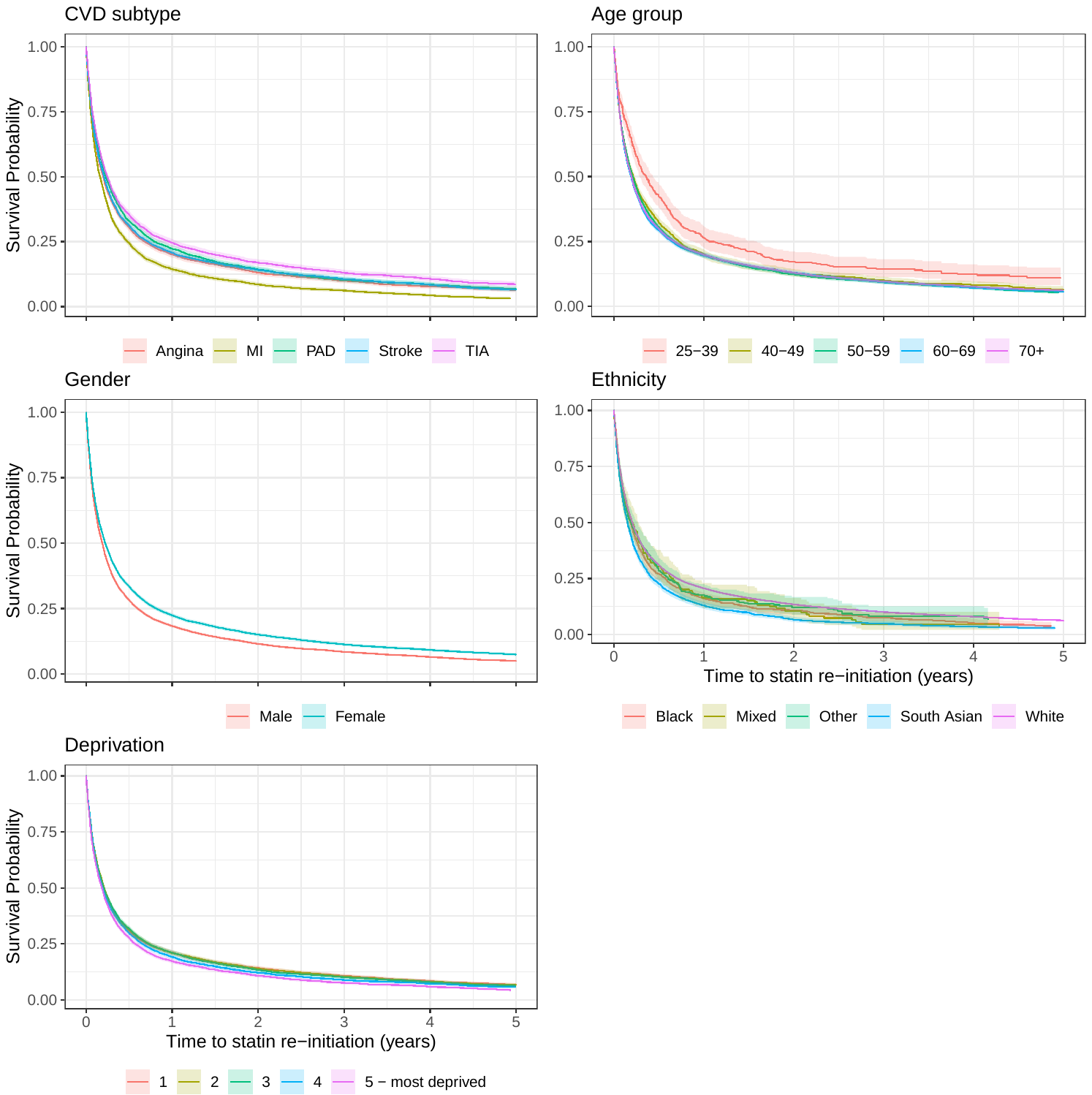

#### Supplementary Table 6: Estimated costs of statins and CVD events among people not prescribed statins scaled to the English population

| **Primary Prevention** | | | | | | | | |
| --- | --- | --- | --- | --- | --- | --- | --- | --- |
| **Statin dose** | **Total population** | **Total quantity** | **Cost per Quantity** | **Total Cost** | **Scaled to England population** | **Weighted average of cost of CVD events** | **Costs saved accounting for statins** | **Costs saved account for statis scaled to English population** |
| Atorvastatin 20mg | 290,246 | 106,012,352 | 0.0420 | £4,450,629.61 | £35,620,809.94 | 2943.87 | £33,179,608.36 | £265,554,455.40 |
| **Secondary Prevention** | | | | | | | | |
| Atorvastatin 80mg | 64,338 | 23,499,455 | 0.0626 | £1,470,448.00 | £11,768,795.27 | 2943.87 | £17,470,390.55 | £139,825,039.46 |

Total population includes individuals in the study who were eligible for but not prescribed statins. The primary prevention cohort includes people with a recorded CVD risk score of 10% or more who were not prescribed statins and the secondary prevention cohort includes people with myocardial infarction, angina, transient ischemic attack and stroke who were not prescribed statins. The cost per quantity is derived from the prescription cost analysis. The total cost is the amount that would be paid using the basic price of statins and the quantity prescribed. The population of England is derived from the 2021 Census of adults aged 25 years and older. The weighted average of cost of CVD events is calculated from the NHS reference costs by dividing the sum of the total cost of CVD events by the sum of activity.

#### Supplementary Figure 13: Adjusted hazard ratios for factors associated with receiving a recorded CVD risk assessment stratified by pre-pandemic and pandemic period

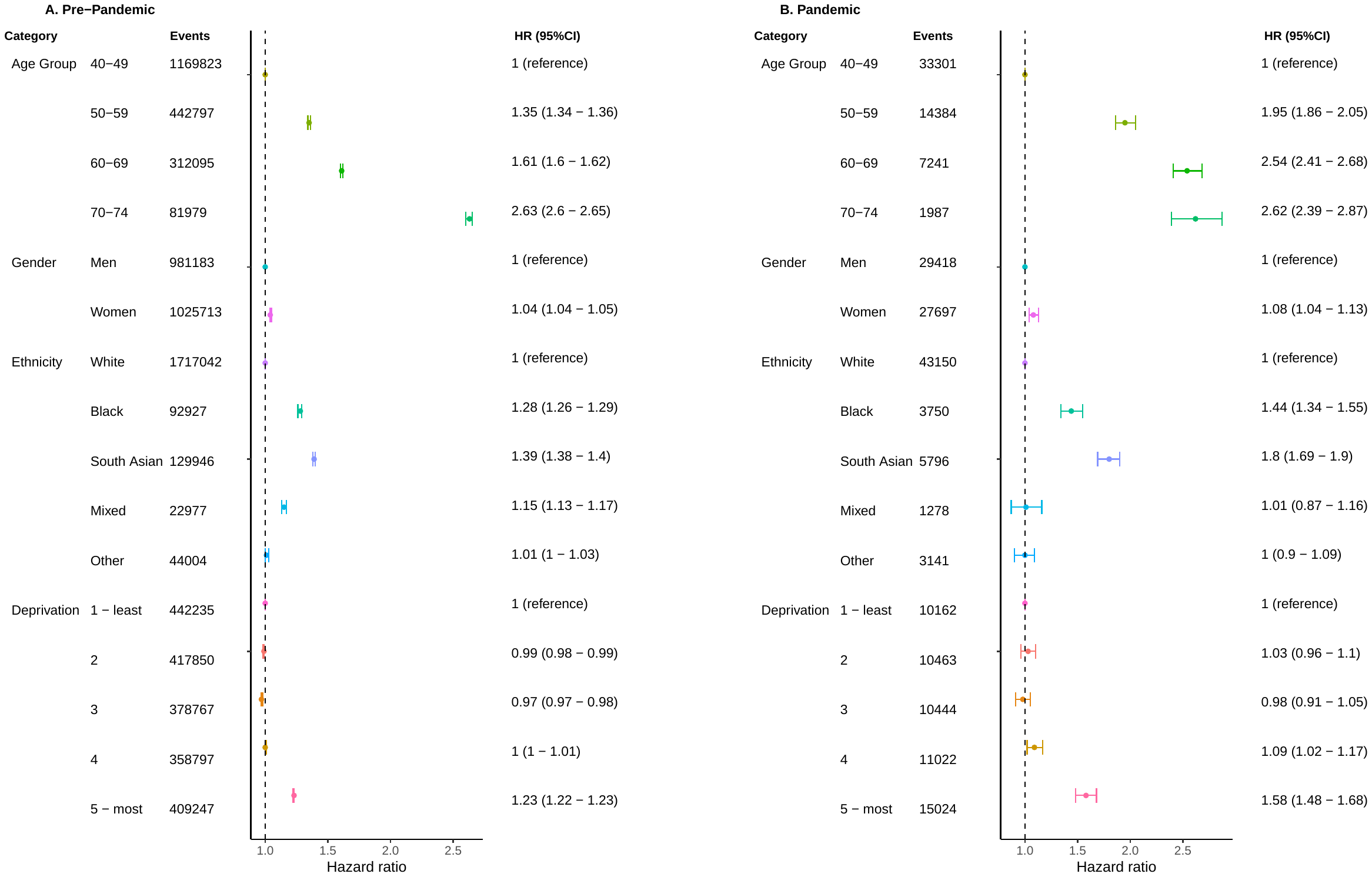

#### Supplementary Figure 14: Crude and adjusted odds ratios for factors associated with statin initiation for primary and secondary prevention stratified by pre-pandemic and pandemic

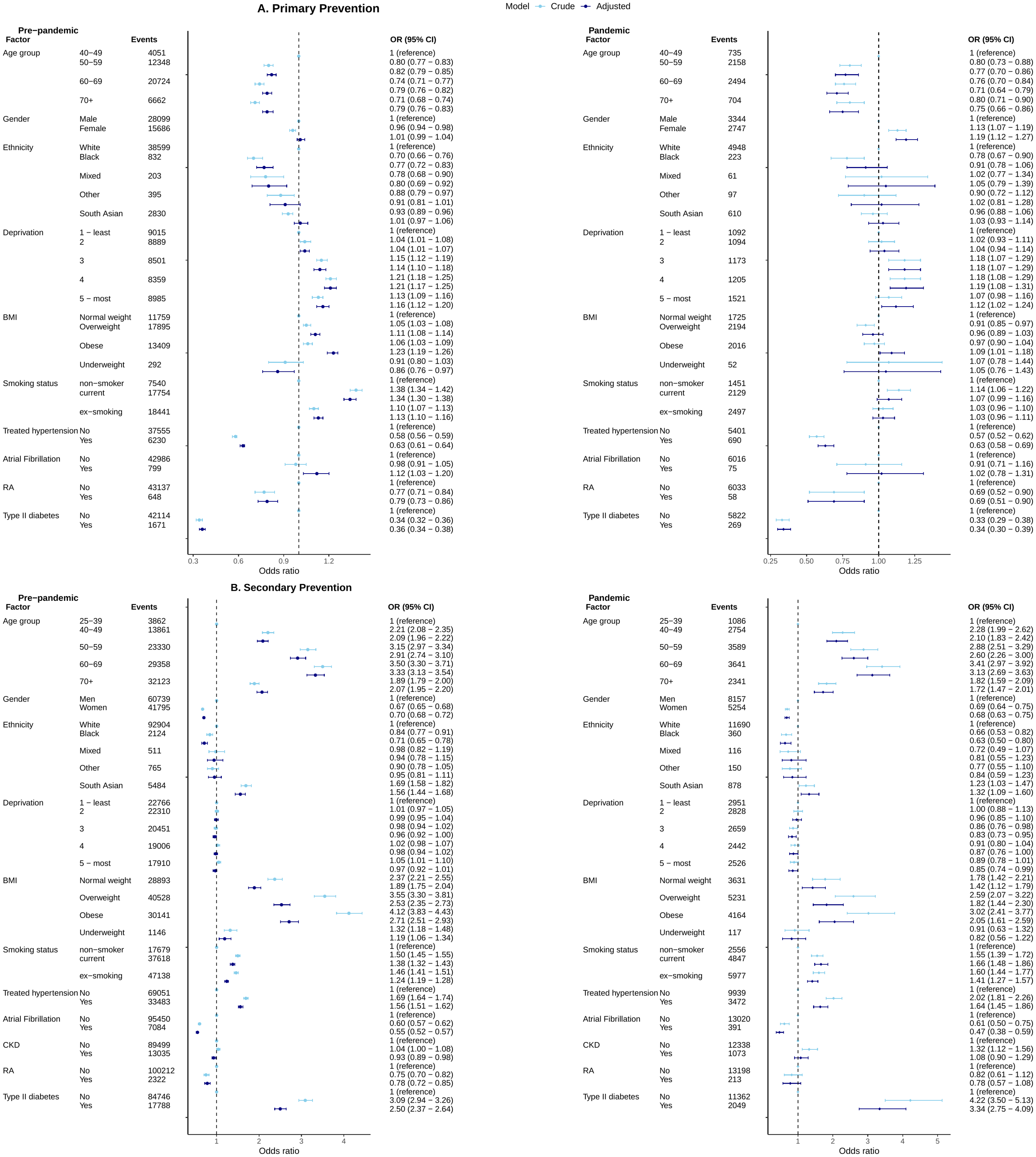

#### Supplementary Figure 15: Adjusted Hazard Ratios for factors associated with statin discontinuation for primary and secondary CVD prevention stratified by pre-pandemic and pandemic period

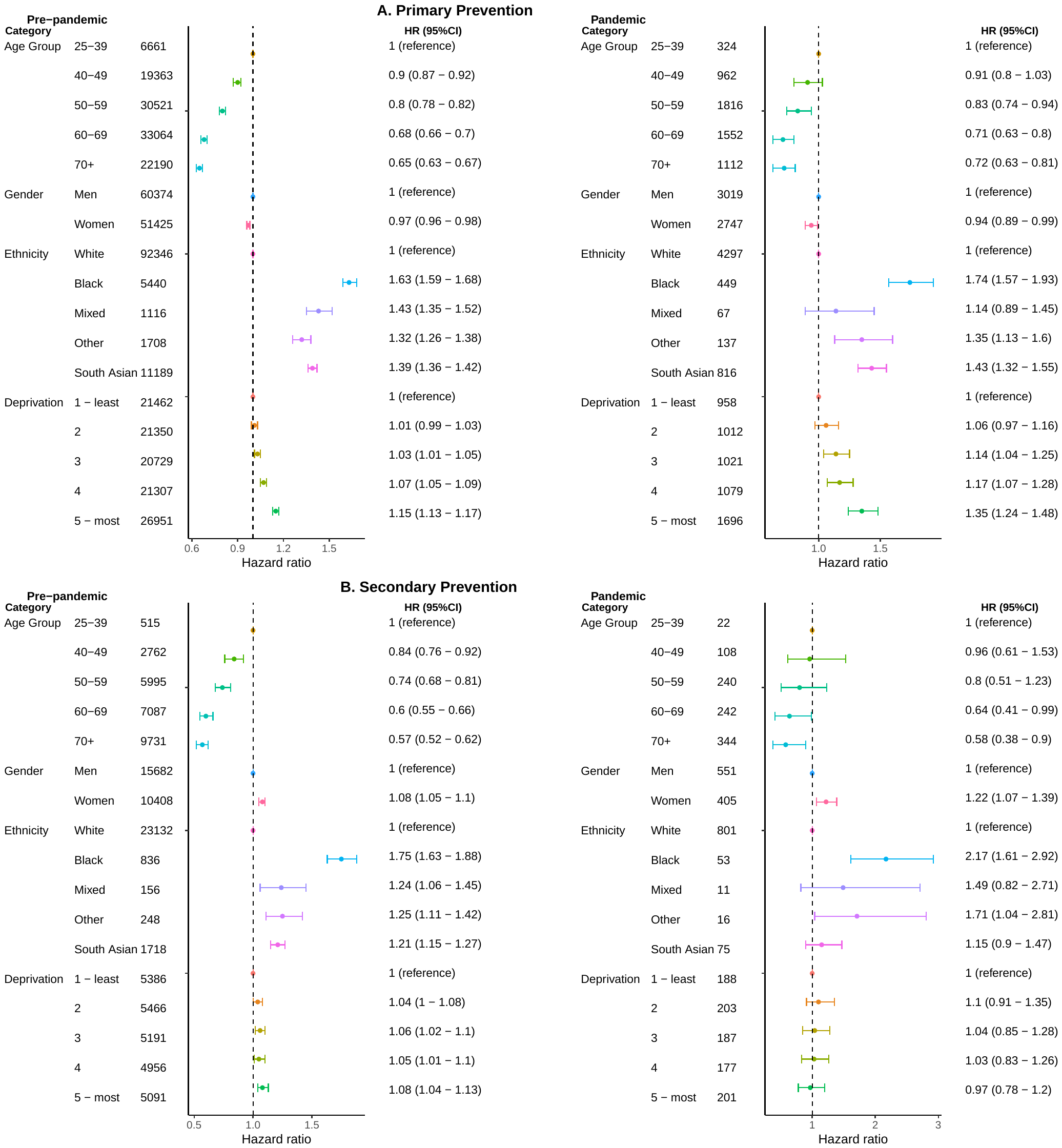

#### Supplementary Figure 16: Adjusted Hazard Ratios for factors associated with statin re-initiation for primary and secondary CVD prevention stratified by pre-pandemic and pandemic period

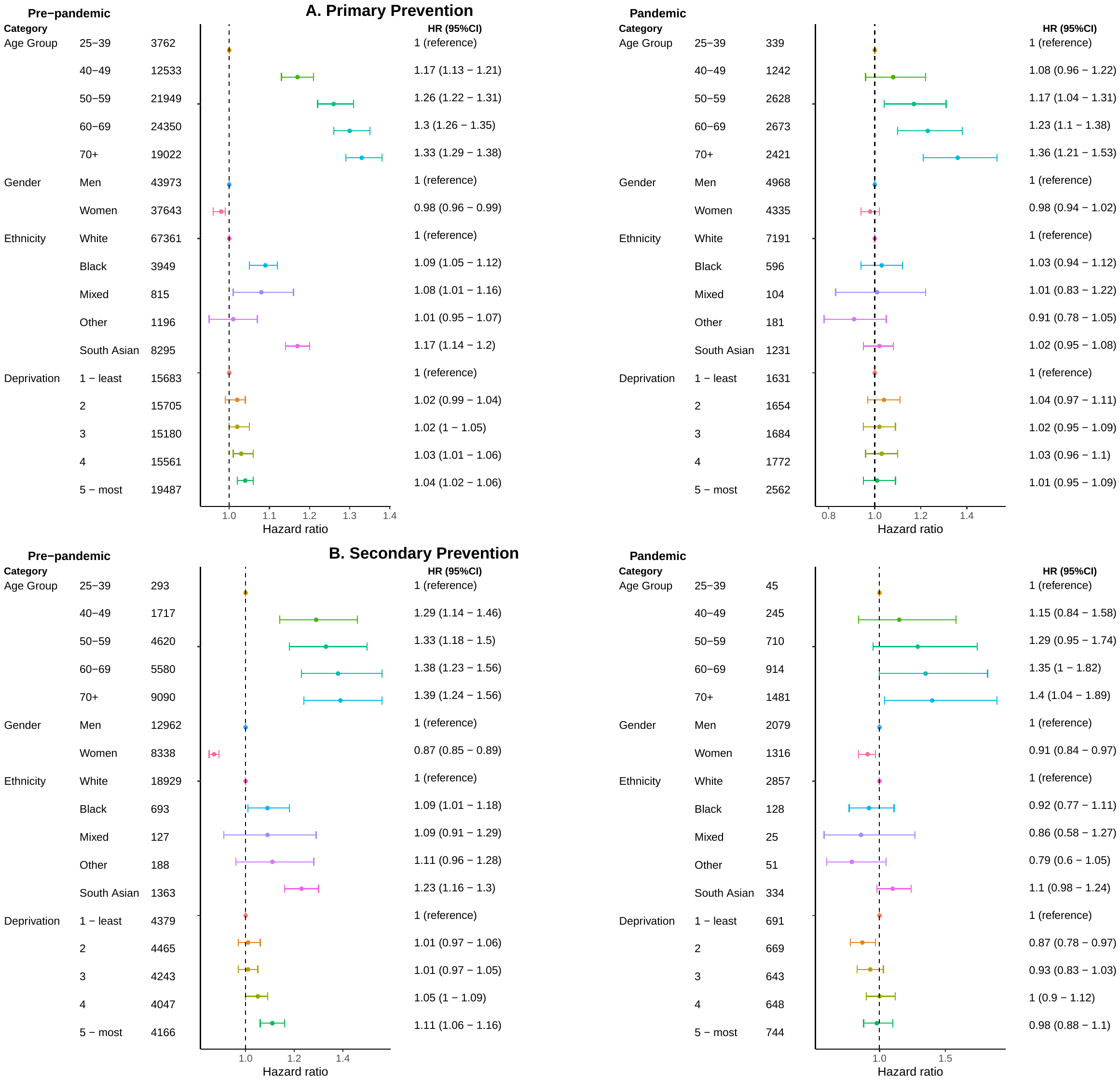

#### Supplementary Figure 17: Crude and adjusted odds ratios for factors associated with statin initiation for primary and secondary prevention, with statin initiators defined as those with a statin prescription within 90 days of above threshold risk score or CVD event

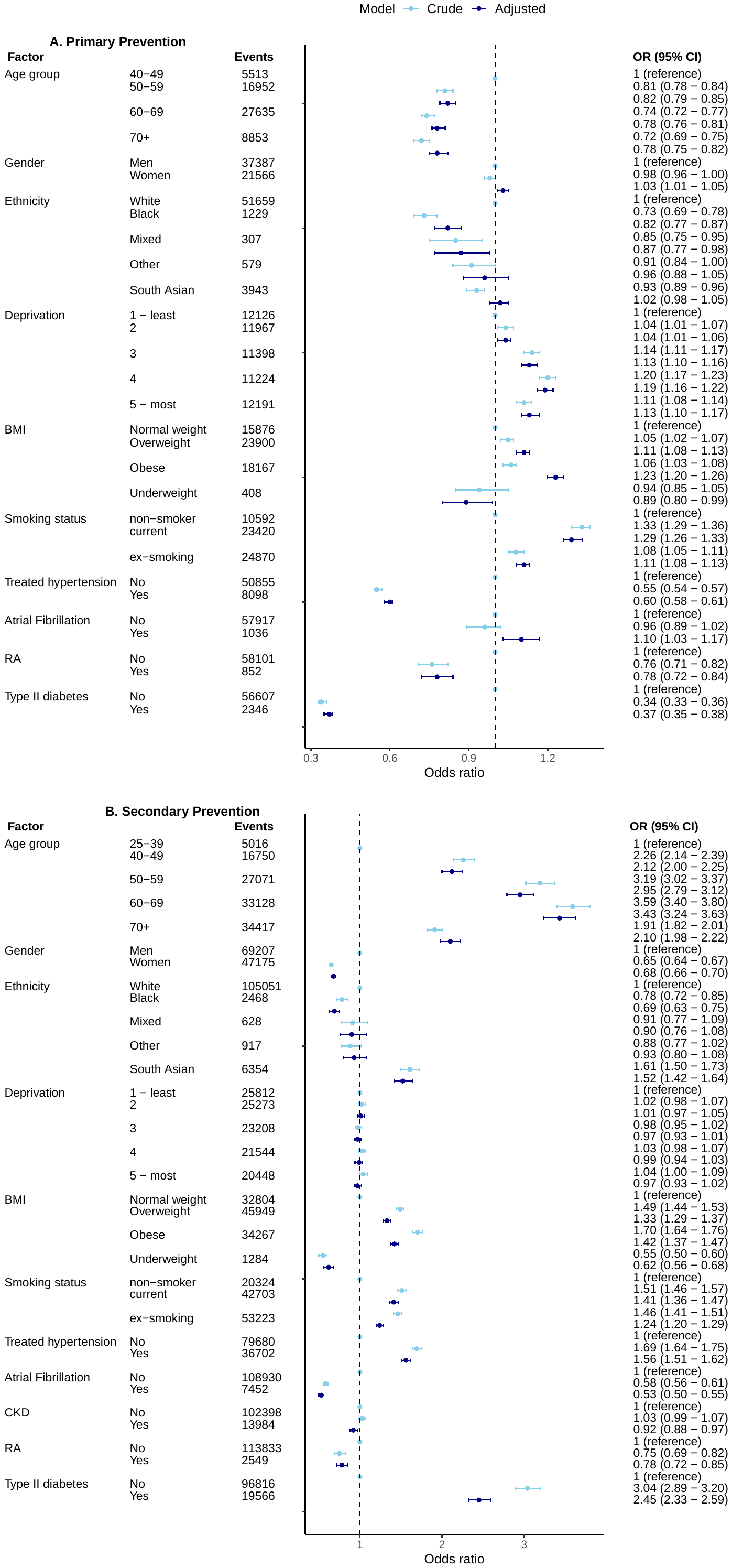

#### Supplementary Figure 18: Adjusted hazard ratios for factors associated with statin discontinuation for primary and secondary prevention, with discontinuation grace period expanded to 180 days

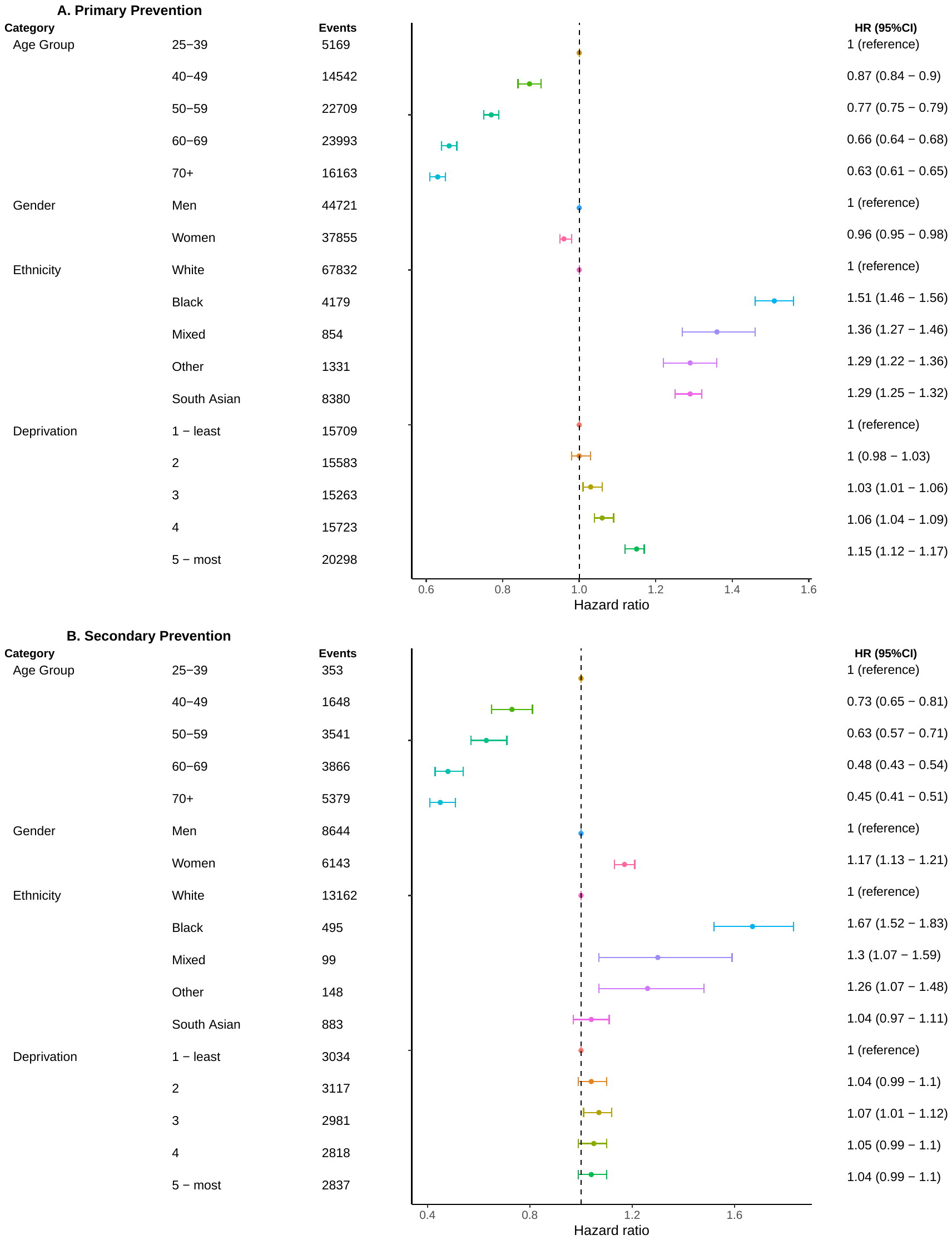

#### Supplementary table 7: Adjusted hazard ratios for the association between demographic variables and risk of receiving a CVD risk assessment, with interaction terms between time and non-proportional factors

| **Demographic factor** | **Adjusted HR (*Time interaction <5 years)** | **95% CI** | ***Adjusted HR (Time interaction 5+ years)** | **95% CI** |
| --- | --- | --- | --- | --- |
| **Age group x Time** |  |  |  |  |
| 40-49 | 1.00 | 1.00 | 1.00 | 1.00 |
| 50-59 | 1.39 | 1.38-1.4 | 0.92 | 0.91-0.93 |
| 60-69 | 1.67 | 1.66-1.68 | 0.87 | 0.86-0.89 |
| 70-74 | 2.68 | 2.65-2.7 | - | - |
| **Gender x Time** |  |  |  |  |
| Men | 1.00 | 1.00 | 1.00 | 1.00 |
| Women | 1.02 | 1.02-1.03 | 1.09 | 1.08-1.1 |
| **Ethnicity x Time** |  |  |  |  |
| White | 1.00 | 1.00 | 1.00 | 1.00 |
| Black | 1.31 | 1.3-1.32 | 0.88 | 0.86-0.9 |
| South Asian | 1.43 | 1.42-1.44 | 0.87 | 0.85-0.88 |
| Mixed | 1.16 | 1.13-1.18 | 0.96 | 0.92-1.01 |
| Other | 1.04 | 1.02-1.05 | 0.9 | 0.87-0.93 |
| **Deprivation x Time** |  |  |  |  |
| 1. Least deprived | 1.00 | 1.00 | 1.00 | 1.00 |
| 2 | 1.01 | 1.00-1.01 | 0.94 | 0.93-0.95 |
| 3 | 1.00 | 0.99-1.01 | 0.91 | 0.9-0.92 |
| 4 | 1.04 | 1.03-1.04 | 0.89 | 0.88-0.91 |
| 5. Most Deprived | 1.30 | 1.29-1.31 | 0.80 | 0.79-0.81 |
| *Start of follow up was the underlying time scale with start of follow up defined as the latest of 1st April 2009, 40th birthday and 12 months after current registration in CPRD. Individuals were followed until the earliest of CVD risk assessment, 75th birthday, CVD event or ineligibility for CVD risk assessment. Evidence of non-proportionality for all covariates was seen using Schoenfelds residual tests and log log plots. HR; Hazard Ratio. | | | | |

#### Supplementary table 8: Adjusted hazard ratios for the association between demographic variables and statin discontinuation for primary prevention, with interaction terms between time and non-proportional factors (age and ethnicity)

| **Demographic factor** | **Adjusted HR** | **95% CI** |
| --- | --- | --- |
| **Age Group x *Time (<6 months)** |  |  |
| 20-39 | 1.00 | 1.00 |
| 40-49 | 0.88 | 0.84-0.91 |
| 50-49 | 0.79 | 0.76-0.82 |
| 60-69 | 0.68 | 0.65-0.7 |
| 70+ | 0.69 | 0.66-0.72 |
| **Gender** |  |  |
| Men | 1.00 | 1.00 |
| Women | 0.97 | 0.95-0.98 |
| **Ethnicity x Time (< 6 months)** |  |  |
| White | 1.00 | 1.00 |
| Black | 1.58 | 1.52-1.64 |
| South Asian | 1.38 | 1.34-1.41 |
| Mixed | 1.36 | 1.26-1.48 |
| Other | 1.35 | 1.27-1.44 |
| **Deprivation** |  |  |
| 1. Least deprived | 1.00 | 1.00 |
| 2 | 1.01 | 0.99-1.03 |
| 3 | 1.04 | 1.02-1.06 |
| 4 | 1.07 | 1.05-1.09 |
| 5. Most Deprived | 1.16 | 1.14-1.18 |
| **Age Group x Time (≥ 6 months)** |  |  |
| 20-39 | 1.00 | 1.00 |
| 40-49 | 1.05 | 0.99-1.11 |
| 50-49 | 1.01 | 0.96-1.07 |
| 60-69 | 1.01 | 0.96-1.07 |
| 70+ | 0.89 | 0.84-0.94 |
| **Ethnicity x Time (≥ 6 months)** |  |  |
| White | 1.00 | 1.00 |
| Black | 1.08 | 1.02-1.14 |
| South Asian | 1.01 | 0.97-1.05 |
| Mixed | 1.06 | 0.94-1.19 |
| Other | 0.93 | 0.85-1.02 |

Date of statin initiation was the time scale used to estimate the association between demographic variables and statin discontinuation using Cox proportional hazards regression models. Evidence of non-proportionality observed for age group and ethnicity using Schoenfelds residual tests and log log plots. HR; Hazard Ratio.

#### Supplementary table 9: Adjusted hazard ratios for the association between demographic variables and statin discontinuation for secondary prevention, with interaction terms between time and non-proportional factors

| **Demographic factor** | **Adjusted HR** | **95% CI** |
| --- | --- | --- |
| **Age Group x Time (<6 years)** |  |  |
| 20-39 | 1 | 1 |
| 40-49 | 0.84 | 0.77-0.93 |
| 50-49 | 0.74 | 0.68-0.81 |
| 60-69 | 0.59 | 0.54-0.65 |
| 70+ | 0.56 | 0.51-0.61 |
| **Gender x Time (<6 years)** |  |  |
| Men | 1 | 1 |
| Women | 1.1 | 1.07-1.13 |
| **Ethnicity x Time (<6 years)** |  |  |
| White | 1 | 1 |
| Black | 1.78 | 1.66-1.91 |
| South Asian | 1.2 | 1.14-1.26 |
| Mixed | 1.24 | 1.06-1.45 |
| Other | 1.26 | 1.11-1.43 |
| **Deprivation** |  |  |
| 1. Least deprived | 1 | 1 |
| 2 | 1.04 | 1-1.08 |
| 3 | 1.06 | 1.02-1.1 |
| 4 | 1.05 | 1.02-1.1 |
| 5. Most Deprived | 1.08 | 1.04-1.12 |
| **Age Group x Time (6≥ years)** |  |  |
| 20-39 | 1 | 1 |
| 40-49 | 1.06 | 0.71-1.57 |
| 50-49 | 1.05 | 0.71-1.53 |
| 60-69 | 1.34 | 0.92-1.96 |
| 70+ | 1.27 | 0.87-1.86 |
| **Gender x Time (6≥ years)** |  |  |
| Men | 1 | 1 |
| Women | 0.76 | 0.69-0.84 |
| **Ethnicity x Time (6≥ years)** |  |  |
| White | 1 | 1 |
| Black | 0.83 | 0.58-1.19 |
| South Asian | 1.07 | 0.88-1.31 |
| Mixed | 0.98 | 0.46-2.09 |
| Other | 1.09 | 0.64-1.83 |

Date of statin initiation was the time scale used to estimate the association between demographic variables and statin discontinuation using Cox proportional hazards regression models. Evidence of non-proportionality observed for age group and ethnicity using Schoenfelds residual tests and log log plots. HR; Hazard Ratio

#### Supplementary table 10: Adjusted hazard ratios for the association between demographic variables and statin re-initiation for primary prevention, with interaction terms between time and non-proportional factors (age group)

| **Demographic factor** | **Adjusted HR** | **95% CI** |
| --- | --- | --- |
| **Age Group x *Time (< 8 years)** |  |  |
| 20-39 | 1.00 | 1.00 |
| 40-49 | 1.16 | 1.12-1.2 |
| 50-49 | 1.26 | 1.21-1.3 |
| 60-69 | 1.29 | 1.25-1.34 |
| 70+ | 1.34 | 1.3-1.39 |
| **Gender** |  |  |
| Men | 1.00 | 1.00 |
| Women | 0.98 | 0.96-0.99 |
| **Ethnicity** |  |  |
| White | 1.00 | 1.00 |
| Black | 1.09 | 1.05-1.12 |
| South Asian | 1.16 | 1.14-1.19 |
| Mixed | 1.08 | 1.01-1.15 |
| Other | 1.00 | 0.95-1.06 |
| **Deprivation** |  |  |
| 1. Least deprived | 1.00 | 1.00 |
| 2 | 1.02 | 1.00-1.04 |
| 3 | 1.02 | 1.00-1.05 |
| 4 | 1.03 | 1.01-1.06 |
| 5. Most Deprived | 1.04 | 1.02-1.06 |
| **Age Group x Time (8 ≥ years)** |  |  |
| 20-39 | 1.00 | 1.00 |
| 40-49 | 1.45 | 1.03-2.05 |
| 50-49 | 1.79 | 1.28-2.5 |
| 60-69 | 1.74 | 1.25-2.43 |
| 70+ | 1.03 | 0.71-1.49 |

*Date of statin discontinuation as the time scale, to estimate hazard ratios (HRs) for the association of age, gender, ethnicity and deprivation with statin discontinuation using Cox proportional hazards regression models. Evidence of non-proportionality observed for age group using Schoenfelds residual tests and log log plots. HR; Hazard Ratio
